# SHERLOCK4HAT: a CRISPR-based tool kit for diagnosis of Human African Trypanosomiasis

**DOI:** 10.1101/2022.03.09.22271543

**Authors:** Núria Sima, Annick Dujeancourt-Henry, Blanca Liliana Perlaza, Marie-Noelle Ungeheuer, Brice Rotureau, Lucy Glover

## Abstract

Elimination of Human African Trypanosomiasis (HAT) requires highly specific and sensitive tools for both diagnostic at point of care and epidemiological surveys. We have adapted SHERLOCK (Specific High-sensitivity Enzymatic Reporter unLOCKing) for the detection of trypanosome nucleic acids. Our SHERLOCK4HAT diagnostic tool kit, using *7SLRNA, TgSGP* and *SRA* targets, distinguishes between *Trypanosoma brucei (T. b*.*) brucei, T. b. gambiense (g) and T. b. rhodesiense* (*r*) without cross-reactivity and with sensitivity between 0.01 and 0.1 parasite/µL. SHERLOCK4HAT can accurately detect a trypanosome infection in cryo-banked patient buffy coats, with 85.1% sensitivity and 98.4% specificity for gHAT, and 100% sensitivity and 94.1% specificity for rHAT. Our SHERLOCK4HAT diagnostic showed 85.6% correlation with a reference standard qPCR in gHAT patients, 96.2% correlation in rHAT patients, discriminates between r/gHAT with 100% accuracy and is compatible with lateral flow assay readout for use at the point of care.

## Introduction

Human African Trypanosomiasis (HAT), or sleeping sickness, is endemic to sub-Saharan countries and without prompt diagnosis and treatment, is usually fatal (*1*). It is caused by a tsetse-borne infection with protist parasites: *Trypanosoma brucei (T. b*.*) gambiense* (gHAT), which represents 85% of the new cases and is endemic to central and western Africa, and *T. b. rhodesiense* (rHAT) which is responsible for the remaining 15% and is found in southern and eastern Africa. Due to both vector control, mass screening and treatment of those infected, the number of gHAT cases is decreasing, and has been maintained below 1,000 new gHAT cases / year since 2018 (*2*). In this context, gHAT has been included in the WHO roadmap to elimination, with zero transmission by 2030 (*2*). Because of the preference of *T. b. rhodesiense* for the animal reservoir and the scarcity of control tools, the complete elimination of rHAT is not considered to be feasible.

The current diagnostic algorithms for gHAT rely on an initial serological test, followed by a parasitological confirmation by direct observation under microscope, which is time consuming, requires trained staff and specialized equipment, and presents a suboptimal diagnostic sensitivity. The reduction in gHAT cases has brought about new challenges, not least that the positive predictive value of any diagnostic test diminishes as the disease burden is reduced. This has been already observed with the gHAT serological tests, the classical Card Agglutination Test for Trypanosomiasis (CATT) and the more recently developed rapid diagnostic tests (RDTs) (review in (*3, 4*)). Moreover, these tests are based on specific surface antigens, which if poorly or not expressed, can lead to missed diagnoses. No serological diagnostic tools are available for rHAT, and diagnosis is still based on clinical manifestations and visual detection of parasites by microscopy. Several molecular amplification tests have been developed for gHAT with promising results (18SrDNA-PCR, TBR-PCR, Tb177bp-qPCR, 18SrDNA-qPCR, SLRNA RT-qPCR, 18S RNA RT-qPCR, RIME-LAMP) (review in (*3, 4*)) but their applications for mass screening are limited due to cost and required infrastructure. Importantly, diagnosis of gHAT is further complicated as there is increasing evidence that the traditional parasitological approaches fail to detect *T. b. gambiense* infections among ‘asymptomatic’ seropositive individuals who are apparently able to control infection to low levels and / or to maintain extravascular parasites, especially in the skin, in the absence of detectable blood parasitemia (*5*). Not only may these individuals contribute to transmission, but they may potentially go on to develop clinical gHAT (*5–7*). Ultimately, specific and high-sensitive tools, suitable for point-of-care (PoC) diagnosis and / or usable in a high-throughput mode in low-income countries are needed.

Adaptation of CRISPR technology towards the development of molecular diagnostics has led to highly sensitive and specific tools for the detection of *Plasmodium* (*8, 9*), Zika, Dengue, SARS-CoV-2, Ebola and *Mycobacterium tuberculosis* to name a few (*10*). SHERLOCK (for Specific High-sensitivity Enzymatic Reporter unLOCKing) is a CRISPR-based approach that relies on the programable collateral nuclease activity of Cas enzymes to identify specific nucleic acid (NA) sequences in samples (*11, 12*). Here, we describe the development of a point-of-care applicable, highly sensitive and specific diagnostic tool that can discriminate between trypanosome species causing g-and rHAT. Our SHERLOCK4HAT tool kit, meets the current WHO recommendations for gHAT diagnostic, and we show its exquisite sensitivity and specificity using RNA from cultured parasites, simulated infections and cryo-banked patient samples.

## Results

### Selection of *Trypanosoma* target regions

The SHERLOCK workflow combines isothermal recombinase polymerase amplification (RPA) with highly specific Cas13-CRISPR RNA target recognition coupled to readout via plate reader (for mass screening) or lateral flow strip (for PoC diagnosis) (Fig. 1A) (*12, 13*). To adapt SHERLOCK for the detection of *T. brucei* sp., we selected gene targets based on the following criteria (i) genes expressed in the human infective form of the parasites, (ii) *T. brucei* species-or subspecies-specific, (iii) degree of conservation between different strains and (iv) few to no single nucleotide polymorphisms (SNPs) (*11, 14*). We assessed several candidate genes, including the superoxide dismutase B1 (*SODB1*) gene (Tb927.11.15910), the component of the peptide recognition particle, *7SLRNA* (Tb927.8.2861), the *T. b. gambiense-*specific glycoprotein gene (*TgSGP*; FN555988.1) and the *T. b. rhodesiense-*specific serum resistance associated gene (*SRA*; AF097331). BLAST analysis revealed that the *SODB1* gene sequence is highly conserved between the *Trypanozoon* subgenus with 99% identity (*T. b. rhodesiense* sequence was not available) and shared certain degree of homology with *T. cruzi* and *Leishmania donovani* genes (77.92% and 76.68% identity, respectively) (data file S1). The *7SLRNA* is highly conserved within the *Trypanozoon* taxa, with 99% identity shared between the *T. brucei* sp., but more distant to the co-endemic species *T. congolense* and *T. vivax* with 86.31% and 79.77% identity respectively (data file S1). *TgSGP* gene is specific to Group 1 *T. b. gambiense* and conserved across isolates in endemic territories (*15–17*). *TgSGP-*like genes, closely related to a possible ancestor *VSG* gene (Tb10.v4.0178), have been identified in other *T. brucei* sspp., with the 5’ region of the genes particularly conserved (*16, 17*). *SRA* is specific to *T. b. rhodesiense* and provides an unbiased identification of the parasite (18–24). Nevertheless, *SRA* is a *VSG*-like gene (*18*) and the first 400 bp share 81% identity with a possible ancestor gene present in the *T. b. brucei* genome (Tb927.9.17380) (*25*). Three *SRA* sequence variants (AF097331, AJ345057, AJ345057) have been identified from field isolates with homology from 97.9-99.7% (*22, 23, 26*) (fig. S1). To develop a pan-*Trypanozoon* SHERLOCK diagnostic, RPA primers and CRISPR RNA guides (crRNAs) were designed to cover the conserved regions of the *7SLRNA* and *SODB1* genes in the *Trypanozoon* subgenus, that are distinct to the other trypanosomatids. For a *T. b. gambiense* and *T. b. rhodesiense* specific SHERLOCK diagnostic, guides were designed to the variable regions of *TgSGP* and *SRA* genes. In an attempt to ensure our tests would be applicable across a wide range of field isolated strains, identified SNPs were considered and guides were designed outside of these regions (tables S1 and S2).

**Fig. 1.**
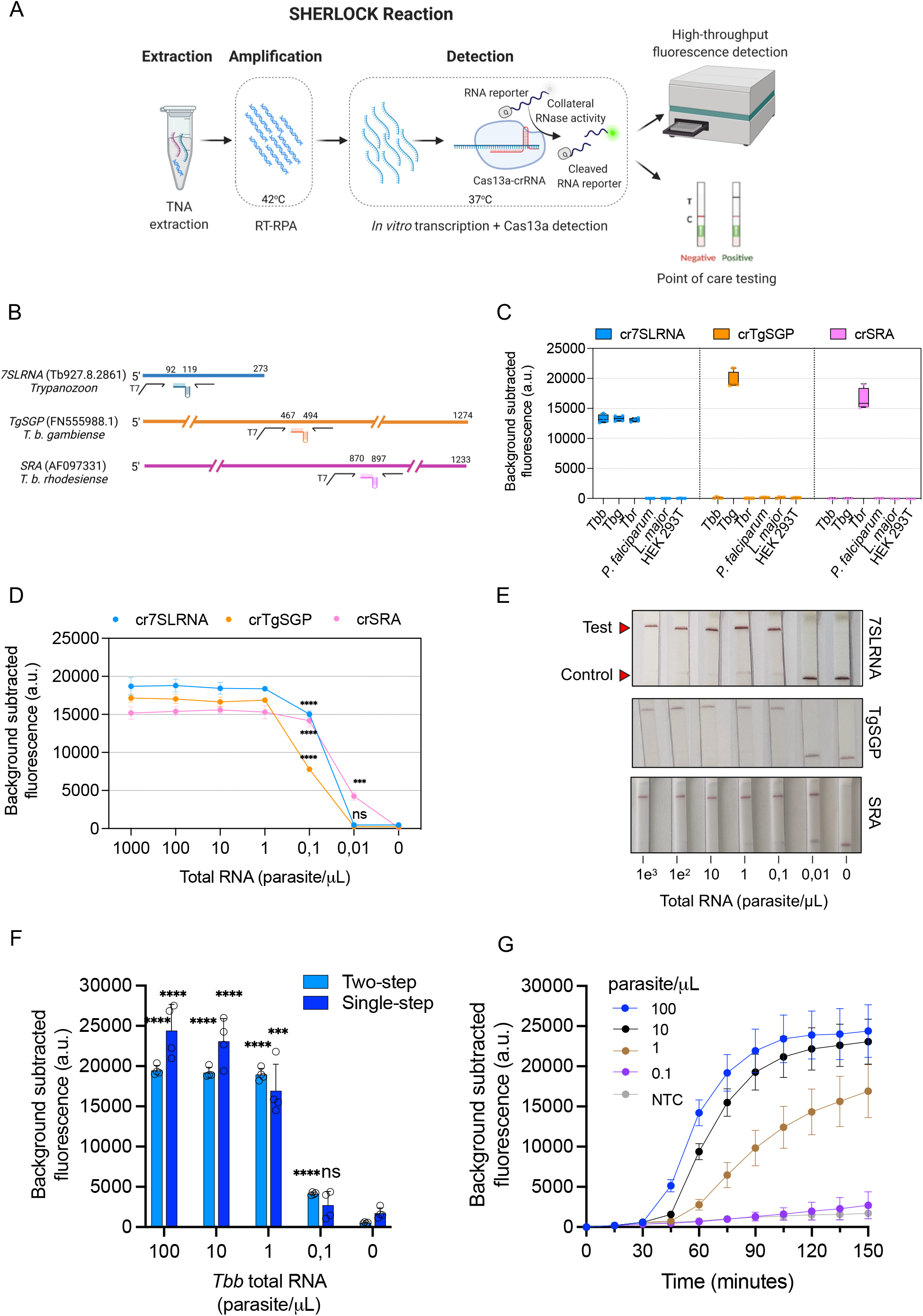
Detection of *Trypanosoma brucei* sspp. RNA with SHERLOCK. **(A), Schematic** overview of the SHERLOCK assay principle. Two-step SHERLOCK reaction is performed after TNA extraction. First, target NA is retro-transcribed and/or amplified during the RT-/RPA reaction at 42°C. Second, the amplified target is *in vitro* transcribed and detected by Cas13a that cuts the RNA reporter upon target activation. Finally, the released reporter can be quantified with a fluorescence plate reader and / or with a LFA, making the methodology suitable for both mass screening and PoC testing. Panel created using BioRender.com. (B), Schematic showing selected target genes, RPA primer pairs and CRISPR guides. (C), Specificity of *7SLRNA, TgSGP* and *SRA* in a two-step SHERLOCK reaction using RNA from *T. b. brucei* Lister 427, *T. b. gambiense* ELIANE strain, *T. b. rhodesiense* EATRO strain, *Plasmodium falciparum, Leishmania major* and Human Embryonic Kidney (HEK) T cells. Fluorescence was measured after 150 mins. Background subtracted fluorescence of 4 technical replicates is plotted as mean +/−standard deviation (SD). a.u., arbitrary units. (D), Limits of detection of the *7SLRNA, TgSGP* and *SRA* targets in two-step SHERLOCK reactions. Dilution series of total RNA extracted from cultured parasites. *T. b. brucei* Lister 427, *T. b. gambiense* ELIANE strain, *T. b. rhodesiense* EATRO strain were used for the *7SLRNA, TgSGP* and *SRA* SHERLOCK reactions, respectively. Fluorescence was measured after 150 mins. Coloured circles represent the mean ± SD of 4 technical replicates. Two-tailed Student’s t test between fluorescence outputs of sample vs. no-template control. *** p<0.001, **** p<0.0001. a.u., arbitrary units. ns, non-significant. (E), Limit of detection of *7SLRNA, TgSGP* and *SRA* in a two-step SHERLOCK reaction with a lateral flow assay (LFA) read-out after 5 mins. (F), Limits of detection of two-step vs. single-step *7SLRNA* SHERLOCK reactions on total RNAs from *T. b. brucei* Lister 427. Fluorescence was measured after 150 mins. Blue bars represent the mean background subtracted fluorescence ± SD of 4 technical replicates shown as open circles. a.u., arbitrary units. (G), Kinetics of the single step reaction in F. Each coloured circle represents the average of 4 technical replicates ± SD. Two-tailed Student’s t test between fluorescence outputs of sample vs. no-template control. *** p<0.001, **** p<0.0001. a.u., arbitrary units. ns, non-significant.

### SHERLOCK distinguishes between the three *T. brucei* subspecies with high sensitivity

We focused on developing a SHERLOCK diagnostic for the two subspecies of *T. brucei* that cause HAT. We screened several RPA primer pairs and crRNA combinations for each of the selected target genes (fig. S2 and table S1 and S3). For *7SLRNA* target, 3 RPA amplicons (Ampl) were combined with 4 crRNA candidates (Ampl 1:crRNAb1-b3, bs, Ampl 2:crRNAb1-b3, Ampl 3:crRNAb1-b3, bs); for *SODB1* target, 6 RPA amplicons and 15 crRNA candidates were studied (Ampl 1:crRNA 1.1-1.3, Ampl 2:crRNA 2.1-2.2, Ampl 3:crRNA 3.3, Ampl 4:crRNA 4.1-4.3, Ampl 5:crRNA 5.1, 5.3, Ampl 6:crRNA 6.1-6.4); 8 RPA amplicons and 23 crRNA were tested for *TgSGP* (Ampl 1:crRNA 1.1-1.3; Ampl 2:crRNA 2.1-2.3; Ampl 3:crRNA 3.1-3.3, Ampl 4:crRNA 4.1-4.3, Ampl 5:crRNA 5.1, 5.3, Ampl 6:crRNA 6.1-6.3, Ampl 7:crRNA 7.1-7.3, Ampl 8:crRNA 8.1-8.3); and 5 RPA amplicons and 14 crRNA were assessed for *SRA* (Ampl 1:crRNA 1.1-1.3, Ampl 2:crRNA 2.1-2.2, Ampl 4:crRNA 4.1-4.3, Ampl 5:crRNA 5.1-5.3, Ampl 8:crRNA 8.1-8.3) (fig. S2A and table S3). A single RPA amplicon-crRNA combination was selected for each gene target based on highest signal-to-noise ratio (fig. S2A) and specificity (fig. S2B) when compared to target recognition in two co-endemic parasite species *Leishmania major* and *Plasmodium falciparum* and to human embryonic kidney (HEK) T cells, as a representation of the host. Based on sensitivity and specificity results, we selected single combination of RPA primers and crRNA sequences for *7SLRNA, TgSGP* and *SRA* for all subsequent analyses (Fig.1B and Table 1). Exquisite specificities were shown for *7SLRNA* as a pan-*Trypanozoon* diagnostic target, and *TgSGP* and *SRA* as species-specific diagnostic targets for *T. b. gambiense* and *T. b. rhodesiense*, respectively (Fig. 1C). The *7SLRNA* SHERLOCK routinely outperformed the *SODB1* SHERLOCK, the second pan-*Trypanozoon* target, hence no further experiments were run with the *SODB1* target. We then wanted to determine the limit of detection (LoD) of each SHERLOCK reaction. The *SRA* SHERLOCK already showed high sensitivity (Fig. 1D), and did not require further optimisation. For the *7SLRNA* and *TgSGP* SHERLOCK reactions, we tested various RPA primer and Magnesium oxalacetate (MgOAc) concentrations (fig. S3), as both can have a direct impact on the amplification rate, and thus on the amplification efficiency. For all subsequent analysis, 480 nM of RPA primer and 14 mM MgOAc were used in the *7SLRNA* SHERLOCK reaction and 240 nM RPA primer and 14 mM MgOAc in the *TgSGP* SHERLOCK reaction (fig. S3). Using input RNA extracted from cultured parasites, the LoD for the *7SLRNA* and *TgSGP* targets was determined to be 0.1 parasite/µL, and the LoD for SRA target was 0.01 parasite/µL, which falls within the range of parasitemia commonly observed in HAT patients (Fig. 1D) (*27, 28*). Using *in vitro* transcribed RNA, the LoD was calculated to be 200 aM (100 molecules/µL) for *7SLRNA* and *TgSGP*, and 20 aM (10 molecules/µL) for *SRA* (fig. S4). This analytical sensitivity is similar to that reported previously for other molecular diagnostics that are subgenus specific (*29–35*) and 10 to 100-fold more sensitive to those reported for subspecies-specific tests (*21, 36, 37*).

**Table 1.**
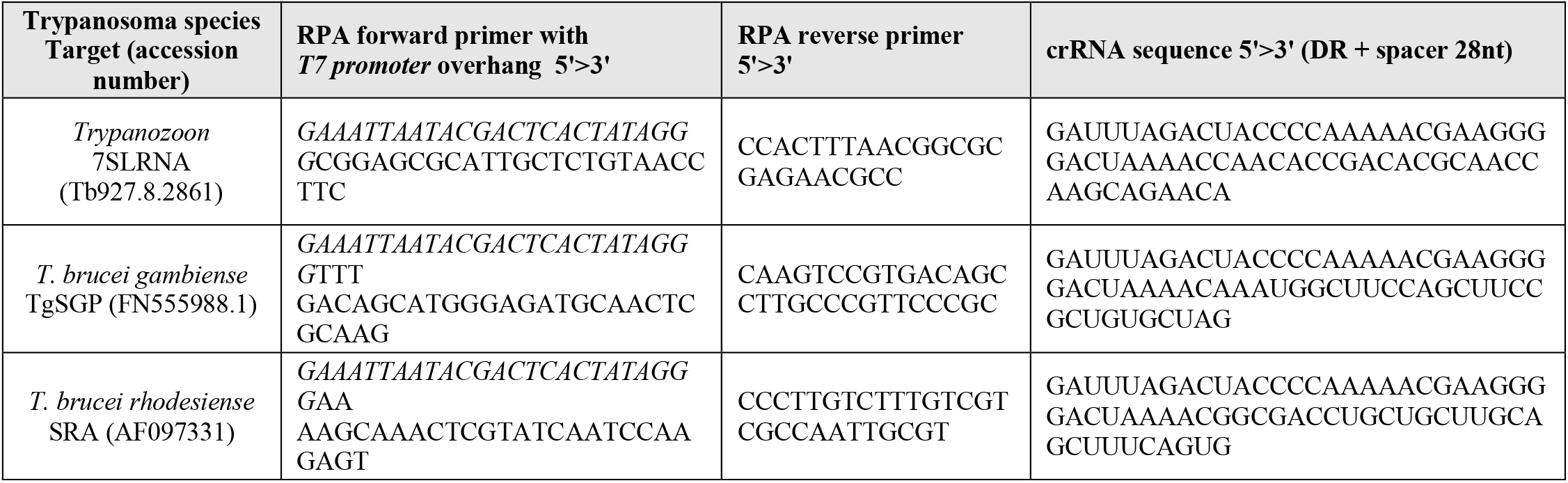
RPA primers and crRNAs selected in this study

#### SHERLOCK can be adapted to a PoC diagnostic use

SHERLOCK is amenable to readout by lateral flow assay (LFA) (*12*). Importantly, using a polyethylene glycol (PEG)-based CRISPR-optimized buffer (provided by Milenia Biotec), we were able to detect the *7SLRNA, TgSGP* and *SRA* SHERLOCK targets with the same respective sensitivities as with the fluorescent readout, but with a reduced background signal as compared to the commercially available LFA buffers, thereby reducing the ambiguity of the readout (Figs. 1E and S5). Our SHERLOCK4HAT diagnostic kit is therefore compatible with use at the PoC. To further optimize SHERLOCK4HAT for PoC use as a one-tube reaction (*8, 38*), we focussed on the *7SLRNA* target and modified the reaction components and conditions (fig. S6). Given that the reverse-transcriptase (RT) and Cas13 activities have different temperature preferences, we evaluated the performance of one-tube reactions at temperatures from 37°C to 42°C, and found that reactions at 37°C had higher signal with reduced sample-to-result time (fig. S6A). An additional consideration for the development of a PoC diagnosis for use in low-income countries is affordability. We therefore tested three RT enzymes from different manufacturers and selected ProtoScript II (NEB) as the most cost-effective reaction with a cost of 2.5 € / reaction (fig. S6B). Given that Cas13a has uridine-cleavage preference (*12, 39*), we compared RNase Alert with a 6U-FAM reporter. The signal intensity obtained with the 6U-FAM reporter was lower and was more prone to spontaneous degradation, as seen with the non-template control reaction (fig. S6C). Thus, we selected RNase Alert as a reporter for an optimized one-tube SHERLOCK diagnostic and used 8 µL of input material (fig. S6D). With these improvements, the *7SLRNA* one-tube SHERLOCK reaction had similar sensitivity than the two-step reaction and detected 1 parasite/µL in less than 1 h (Fig. 1, F and G).

#### The SHERLOCK4HAT diagnostic kit can accurately detect a trypanosome infection across multiple regions and over extended periods of time

Genetic variability between field isolates can potentially lead to false negatives with molecular diagnostic. Therefore, the development of a robust diagnostic hinges upon the ability to detect all parasite strains or variants. To demonstrate the robust specificity of our SHERLOCK4HAT diagnostic kit, we analyzed total RNAs from 57 *Trypanozoon* strains, isolated from their host over the course of 50 years and maintained at the Institute of Tropical Medicine (ITM, Antwerp, Belgium) (*40*) (table S4). Using our two-step SHERLOCK assay, all samples were positive for *7SLRNA*, confirming *7SLRNA* SHERLOCK as a pan-*Trypanozoon* diagnostic and epidemiological tool (Fig. 2 and table S4). Within this set, the 46 *T. b. gambiense* Group 1 mammalian stage isolates tested positive for *TgSGP*, and 3 out of 4 *T. b. gambiense* Group 1 insect stage isolates were negative, as expected since *TgSGP* is only expressed in the mammalian stage of the parasite (*15*). The single *T. b. gambiense* Group 1 insect stage isolate (MHOM/CI/91/SIQUE1623) that was positive for *TgSGP*, may have retained low level expression of the gene. The 7 non-Group 1 *T. b. gambiense* strains tested negative for *TgSGP*, including the T. *b. gambiense* Group 2 sample (Fig. 2, and table S4), thus confirming the diagnostic specificity of the *TgSGP* SHERLOCK for *T. b. gambiense* Group 1. The two *T. b. rhodesiense* strains included in the collection were positive for *SRA*, and 54 of the 55 non-*T. b. rhodesiense* strains tested negative for *SRA* (Fig. 2 and table S4). A single isolate, AnTat 22.1, classified as *T. b. gambiense* Group 1 was positive for both *TgSGP* SHERLOCK and *SRA* SHERLOCK (table S4). Sequence analysis showed that the *SRA* SHERLOCK target amplicon shared 83.3% identity with a *VSG* (Tbb1125VSG-4336, accession number: KX700900) that was expressed in this sample (fig. S7A). There are 7 nucleotide mismatches between the *SRA* guide and the homologous region in the *VSG* sequence (fig. S7B), and this is the most likely source of the cross-reactivity. In spite of the 1.8% cross-reactivity observed with the *SRA* SHERLOCK within the group of samples analyzed, 100% of the strains were detected with the corresponding test, confirming the use of *TgSGP* SHERLOCK and *SRA* SHERLOCK for diagnosis across endemic regions.

**Fig. 2.**
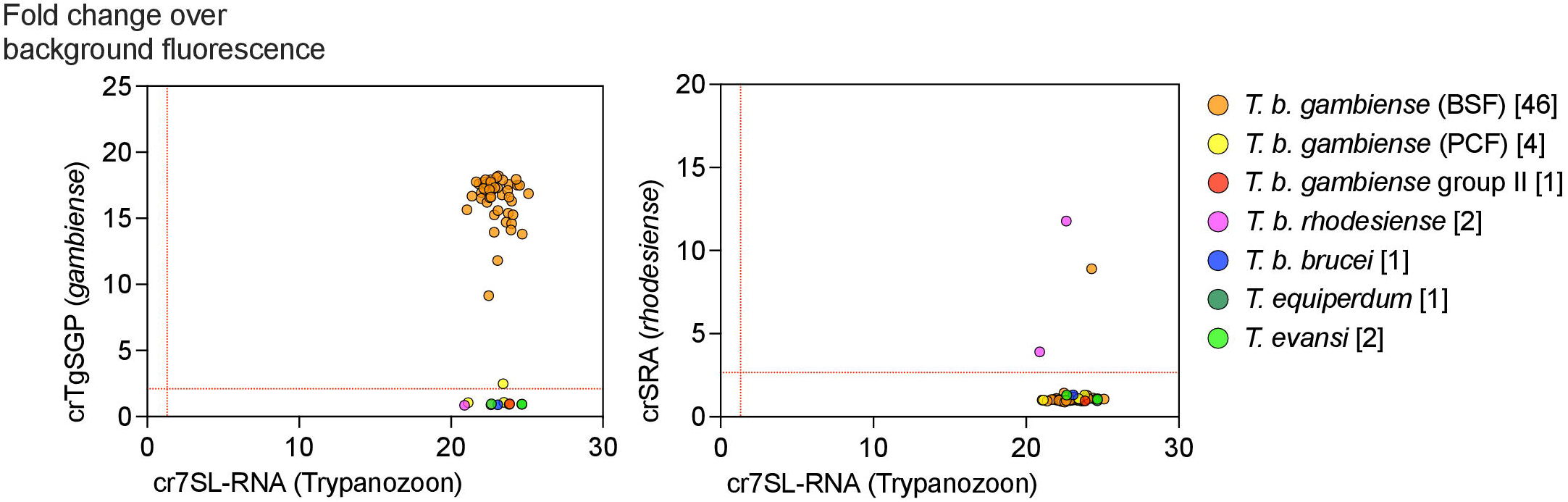
Validation of SHERLOCK4HAT using field isolated samples. SHERLOCK detection of *7SLRNA, TgSGP* or *SRA* targets using RNA extracted from field isolated trypanosome strains. *TgSGP* (left panel) and *SRA* (right panel) target readouts are plotted against *7SLRNA* readouts in fold change over background fluorescence. Each dot represents the average readout of 4 technical replicates. The thresholds for each target (red lines) were determined using ROC curve analyses of positive and negative sample data. In brackets, after the name of the species, number of strains analysed.

#### SHERLOCK4HAT detects trypanosomes in dried blood spots, whole blood and buffy coat

There is a critical need for highly sensitive and specific molecular detection tools that can be use in a high-throughput format in the context of gHAT post-elimination phase. Adapted strategies will be adopted to monitor for potential residual transmission and these tests will be performed in regional reference centres. Methods to capture individual samples, such as dried blood spots (DBSs) would allow easy collection in the field and safe transport and storage back to a lab. SHERLOCK can detect both DNA and RNA, therefore, working with total nucleic acid (TNA) instead of RNA alone can increase the sensitivity. However, for use as a test of cure it is important to work with RNA only, since trypanosome DNA has been detected in the host up to 2 years after cure (*35*). To optimize the *7SLRNA* SHERLOCK for epidemiological surveys, we compared three methods of TNA extraction from DBS using non-infected sheep blood spiked with cultured *T. brucei* parasites spotted on Whatman 903™ Cards. Our *7SLRNA* SHERLOCK was able to detect 100 parasites/µL using a RNeasy kit (Qiagen) and 10 parasites/µL with the NucleoSpin Triprep kit (Macherey-Nagel) (fig. S8A). We saw consistently greater sensitivity with the NucleoSpin Triprep kit, and therefore used it for subsequent extractions from DBSs. Mass screening campaigns are expected to result in a high volume of samples that require subsequent processing, thus, an automated system that minimizes the hands-on time in the extraction process and the cross-contamination between samples is preferred. We compared the performance of two different kits from the automated, paramagnetic beads-based system Maxwell RSC (Promega) and the manual column-based system from Qiagen (fig. S8, B and C). Maxwell RSC DNA blood kit was more efficient than Maxwell RSC RNA kit for TNA extraction using simulated infected samples and showed no cross-contamination, in contrast to the manual column-base kit (fig. S8, B and C). Using simulated human infections (un-infected human blood spiked with *T. brucei* parasites), we compared the performance of the *7SLRNA* SHERLOCK using DBS, whole blood and buffy coat. *7SLRNA* SHERLOCK detected trypanosome TNAs equivalent to 1 parasite/µL in the three types of samples (Fig. 3A), which is in line with the analytical sensitivity reported previously with the M18S-qPCR in DBS (*31*). To determine the *7SLRNA* SHERLOCK analytical sensitivity, we analyzed 3 independent dilution series of simulated infections in whole blood and buffy coat samples and estimated the LoD to be the lowest concentration where 3 out of 3 samples tested positive (Fig. 3, B and D). Here, the resulting analytical sensitivity was determined to be 10 parasites/µL in whole blood and 1 parasite/µL in buffy coat. This is consistent with thet increased sensitivity seen with the mini-anion exchange centrifugation technique (mAECT) when buffy coat is analyzed instead of whole blood (*27*). We further resolved the analytical sensitivity of the *7SLRNA* SHERLOCK with buffy coat by analysing 20 replicates of simulated infected samples at 0.66X, 1X or 1.5X a 1 parasite/µL parasitemia. We confirmed the *7SLRNA* SHERLOCK buffy coat LoD to be 1 parasite/µL in 95% of the samples detected (Fig. 3, C and D). Further improvement of the extraction methods will be required to increase the analytical sensitivity, since the LoD of SHERLOCK tests with RNA from cultured parasites was more sensitive (Fig. 1D).

**Fig. 3.**
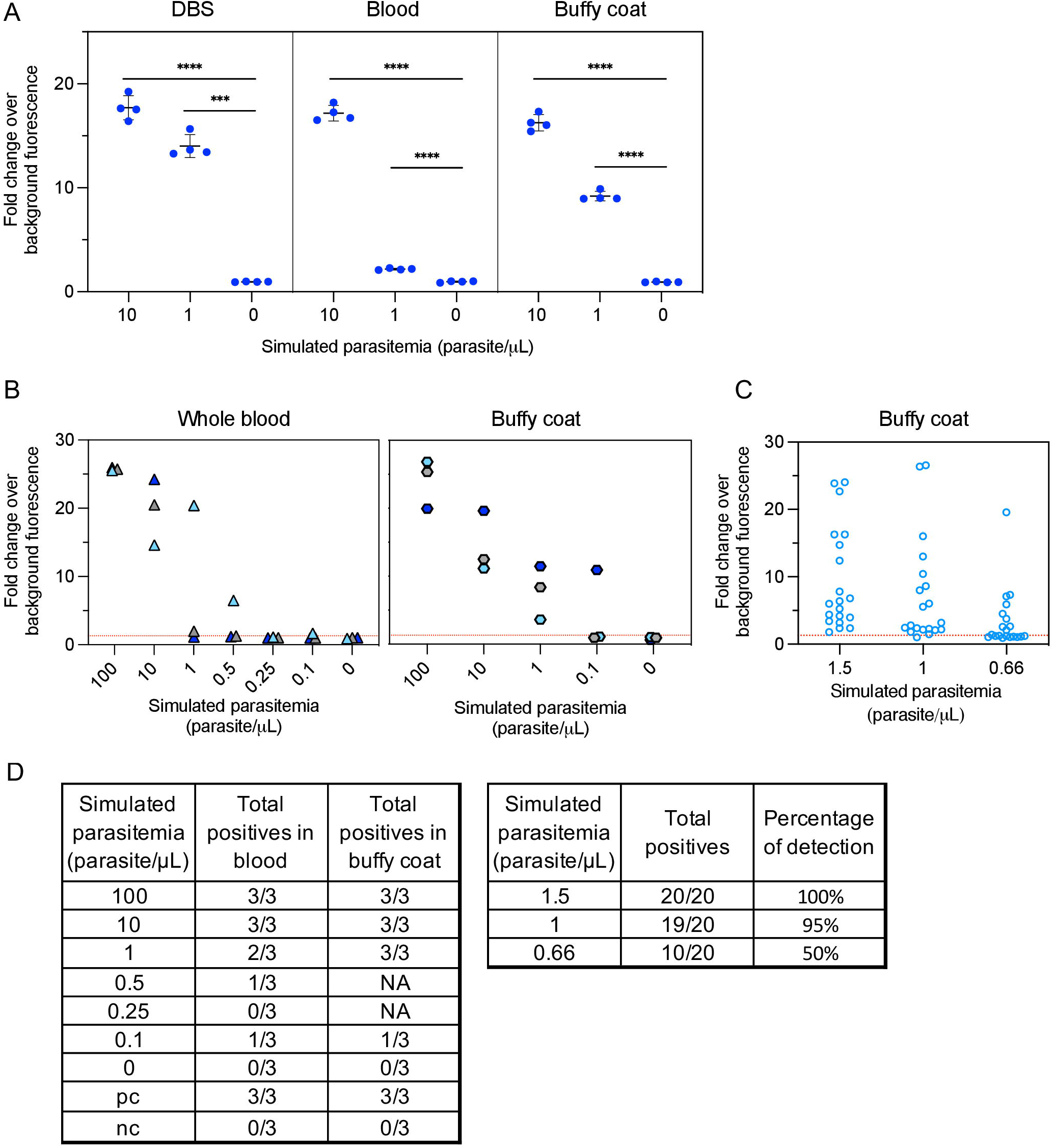
Performance of SHERLOCK4HAT on dried blood spots, whole blood and buffy coat. **(A)**, A comparison of the performance of the *7SLRNA* SHERLOCK on trypanosome RNA extracted from dried blood spots (DBS), whole blood and buffy coat. All experiments were done in 4 replicates from a single pool of simulated infected blood. Fold changes over background fluorescence were plotted as mean +/−SD. Student’s two-tailed t test between readout of sample vs. no-template control. *** p<0.001, **** p<0.0001. **(B)**, Samples with known parasitemia were used to assess the analytical sensitivity of the *7SLRNA* SHERLOCK. Three replicates of each dilution were tested. The tentative limit of detection (LoD) was the lowest concentration where 3/3 replicates were positive for the test. The detection threshold (red line) was determined using ROC curve analyses of positive and negative sample data. **(C)**, The LoD was confirmed by using samples at 0.66X, 1X and 1.5X the estimated LoD concentration of the buffy coat only. The experiment was done on 20 replicates and the LoD was determined to be the concentration at which 95% of the samples were positive for the test. The detection threshold (red line) was determined using ROC curve analyses of positive and negative sample data. **(D)**, Tables summarizing the results in panels B and C. Performance near the LoD of *7SLRNA* SHERLOCK with whole blood and buffy coat samples.

#### Validation of the SHERLOCK4HAT diagnostic kit using clinical samples

To validate SHERLOCK4HAT as a diagnostic tool kit, we used samples obtained from the WHO HAT specimen biobank (*41*): 98 buffy coat samples from patients with confirmed gHAT; 48 buffy coat samples from gHAT endemic regions, but negative for gHAT, to act as controls; 19 buffy coat samples from patients with confirmed rHAT, and 20 buffy coat samples from rHAT endemic regions, but negative for rHAT, as further negative controls. Additionally, we analyzed 14 buffy coat samples from un-infected donors from non-endemic regions. As a positive control for TNA extraction to validate negative SHERLOCK results in clinical samples, i.e. to ensure that no SHERLOCK inhibitors were remaining in the sample, we designed an additional SHERLOCK assay that targeted the human *RNase P* gene and validated its performance using RNA from cultured human cells and parasites (fig. S9). All samples were maintained at −80°C from collection until delivery by the WHO HAT specimen biobank, and all the samples tested here were more than 10 years old and stored without any preservative (*41*). Therefore, the likelihood of TNA deterioration was high, especially for RNA. As an additional control, we ran a *Tb177 bp repeat* qPCR (*32*) in parallel. Trypanosome DNA was detected by qPCR in 47 out of 98 confirmed gHAT samples, suggesting deterioration of the TNAs in most of the samples (Fig. 4, A and B and data file S2). Notably, the *7SLRNA* SHERLOCK detected 55 out of 98 confirmed gHAT samples, showing a higher sensitivity as compared to the reference qPCR test. The concordance between the two assays was 85.6%, with 40 out of 47 qPCR positive samples and 97 out of 113 qPCR negative samples, positive and negative for SHERLOCK respectively (Fig. 4, A and B and data file S2). Importantly, 15 of the 16 qPCR negative samples that tested positive for SHERLOCK were part of the originally confirmed gHAT cohort, revealing our SHERLOCK diagnostic to be more sensitive than the qPCR test (Fig. 4, A and B and data file S2). None of the gHAT endemic negative control samples tested positive, and one out of 14 non-endemic negative control samples was positive using the *7SLRNA* SHERLOCK with an overall specificity of 98.4%. The sensitivity of the *7SLRNA* SHERLOCK was 85.1 % for gHAT, based on the qPCR positive samples (Fig. 4B). The *TgSGP* SHERLOCK detected 42.5% of the qPCR positive samples with 88.7% specificity (data file S2 and table S5). These differences in sensitivity between the spp-specific and the pan-*Trypanozoon* SHERLOCK are most probably due to a selective degradation of the target RNAs. All 19 confirmed rHAT patient samples tested positive for *7SLRNA* SHERLOCK with 100% sensitivity and 94.1% specificity (Fig. 4, C and D and data file S2). To note, the impact of TNA deterioration in these samples may be lower due to the higher parasitemia in *T. b. rhodesiense* infections (*1*), seen here by the lower qPCR Ct values in rHAT samples compared to those in gHAT samples (Fig. 4, A and C and data file S2). The *SRA* SHERLOCK detected 79% of the qPCR positive samples with 100% specificity (data file S2 and table S5). The combination of results from the *7SLRNA, TgSGP* and *SRA* SHERLOCK diagnostics suggests an increased sensitivity of the single tests, but slightly reduced specificity (Fig. 4, B and D and table S5). No correlation between the stages of the disease and the qPCR Ct values or the SHERLOCK fluorescence readouts in gHAT patients was observed (Fig. 4A and data file S2). Results of the 19 confirmed rHAT samples with *TgSGP* SHERLOCK were negative. Likewise, 20 selected confirmed gHAT samples were all negative when run using the *SRA* SHERLOCK diagnosis protocol (Fig. 4E). Thus, the *TgSGP* and *SRA* SHERLOCK in patient samples accurately discriminate between *T. b. gambiense* and *T. b. rhodesiense* infections.

**Fig. 4.**
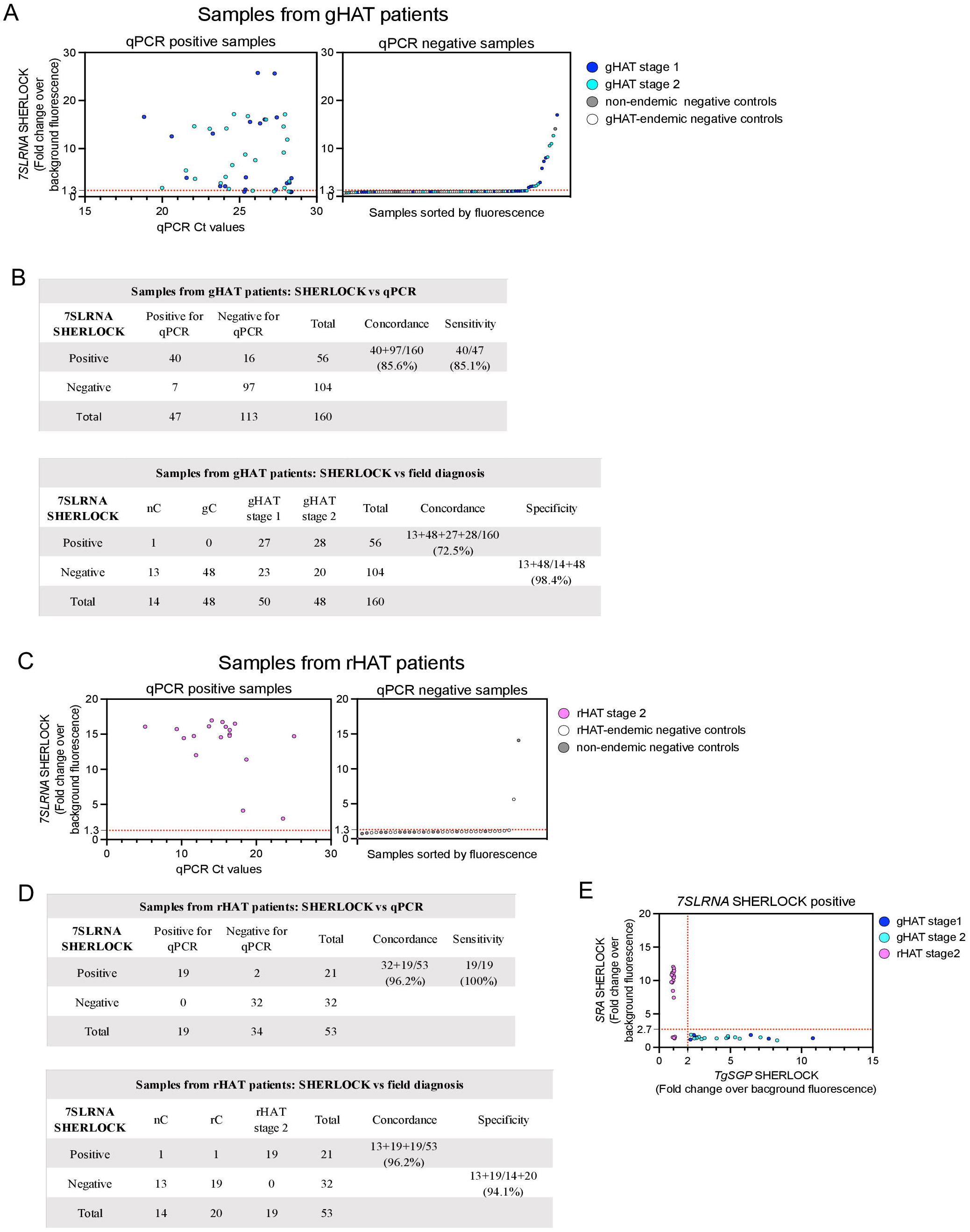
Validation of SHERLOCK4HAT diagnostic using bio-banked clinical samples. **(A)**, *7SLRNA* SHERLOCK analysis of 160 gHAT qPCR positive (left) and negative (right) buffy coat samples obtained from human subjects in gHAT endemic and non-endemic regions. Each dot indicates one sample analysed by both *7SLRNA* SHERLOCK and *Tb177bp-repeats*-qPCR. The dashed red line is the threshold above which samples were considered positive for SHERLOCK. The dot colours indicate the classification according to the original diagnosis in the field. **(B)**, Concordance tables, sensitivity and specificity of *7SLRNA* SHERLOCK for detection of trypanosome NA in buffy coat samples from individuals in gHAT endemic and non-endemic regions. qPCR indicates samples that were analysed by *Tb177bp-repeats-*qPCR. nC, negative controls from non-endemic regions, gC, gHAT negative endemic controls. **(C)**, *7SLRNA* SHERLOCK analysis of 53 rHAT qPCR positive (left) and negative (right) buffy coat samples obtained from human subjects in rHAT endemic and non-endemic regions. Each dot indicates one sample analysed by both *7SLRNA* SHERLOCK and *Tb177bp-repeats*-qPCR. The dashed red line is the threshold above which samples were considered positive for SHERLOCK. The dot colours indicate the classification according to the original diagnostic in the field. **(D)**, Concordance tables, sensitivity and specificity of *7SLRNA* SHERLOCK for detection of trypanosome NA in buffy coat samples from individuals in rHAT endemic and non-endemic regions. qPCR indicates samples that were analysed by *Tb177bp-repeats-*qPCR. nC, non-endemic negative controls, rC, rHAT endemic negative controls. **(E)**, *TgSGP* and *SRA* SHERLOCK discriminate between *T. b. gambiense* and *T. b. rhodesiense* NA in patients. All gHAT and rHAT patient samples positive for *7SLRNA* were plotted according to their *TgSGP* and *SRA* SHERLOCK results. The thresholds for each target (red lines) were determined using ROC curve analyses of positive and negative sample data.

## Discussion

Here we described the development of a new molecular detection tool for HAT diagnosis adapted for use at a PoC and for HAT epidemiological surveillance. Our SHERLOCK4HAT diagnostic can distinguish between the three *T. brucei* subspecies using a pan-*Trypanozoon, gambiense*-specific, or *rhodesiense*-specific targets. Although our subspecies-specific targets use *TgSGP* and *SRA*, which are related to *VSG* genes, we do not see cross reactivity. In fact, in spite of the degree of DNA sequence homology shared between *SRA* and *VSG* variants, we saw no false positives, confirming that the selected target meets the specificity requirements for rHAT diagnosis.

As an RNA based diagnostic, SHERLOCK4HAT is a highly sensitive detection method for on-going infections with a simple set up. We show that the analytical sensitivity of SHERLOCK4HAT for the *Trypanozoon* target is 0.1 parasite/µL (100 parasites/mL), which is comparable to several molecular techniques for detection of *Trypanozoon* taxa (TBR-PCR/qPCR and 18S-PCR, 50-100 parasites/mL). However, implementation of these techniques is limited by the need for sophisticated equipment *(29, 31, 32, 42)*. An additional advantage of SHERLOCK4HAT is the single temperature isothermal RPA amplification coupled to a Cas13 detection, making our method more adapted to the low-income countries where the disease is endemic. Other isothermal approaches have been developed with similar sensitivities to SHERLOCK (LAMP-100 parasites/mL, NASBA-10 parasites/mL) (*43, 44*), but significant infrastructure costs have limited their implementation in control programs. Our subspecies-specific SHERLOCK4HAT diagnostics using a *TgSGP* or *SRA* target are 10 to 100-fold more sensitive than that the current subspecies-specific diagnostics using PCR/qPCR (*21, 36, 45*) and show no overlap between the signal in positive and negative samples, resolving any ambiguity seen in PCR and qPCR. In fact, our results indicated that SHERLOCK4HAT can unequivocally discriminate between *TgSGP* and *SRA*, and therefore diagnose of *T. b. gambiense* and *T. b. rhodesiense* infections.

The current gHAT field-applicable diagnostic algorithms are based on antibody detection in patient blood (CATT and RDTs) with confirmation of seropositive cases by parasitological observation (*1, 4*). These methods present some limitations: (i) false-negative results if the *VSG* variants detected by the test are poorly or not expressed (*46*), (ii) reduced specificity (*1, 4*), and (iii) relatively high cost (RDTs), significant workload (CATT on serial plasma dilution, time at microscope) and need for specialized staff and equipment (especially for parasitological confirmation). For rHAT, no field-applicable diagnostic methods exist. SHERLOCK4HAT overcomes these limitations. We have shown that SHERLOCK4HAT detection is not limited by geography or time after sampling, it is easy to set up, does not require sophisticated equipment and it is adapted for high-throughput applications (fluorescence readout) or individual testing (LFA), making it versatile for both surveillance at reference centers and PoC testing. The SHERLOCK4HAT diagnostic can be run in 1 h 30 min for a one-step reaction (at 4.5 € if coupled to a commercial LFA), or 2 h 30 min for a two-step reaction (at 2.8 €), and these costs would be notably reduced with an in-house manufactured strip for LFA.

One limitation for SHERLOCK4HAT, as for any molecular diagnostic method, is the NA extraction step. Several extraction methods coupled to a CRISPR-based diagnostic reaction have been published (*8, 47, 48*), but remained to be tested in the context of HAT diagnostics. For high-throughput surveillance using SHERLOCK4HAT, automated NA extraction systems can be implemented with higher reproducibility, reduced hand-on time and no cross-contamination. Manual extraction methods, although more time consuming, showed an increased analytical sensitivity which is consistent with previous studies (*49*). As an RNA-based diagnostic, SHERLCOCK is limited by the increased sensitivity of RNA to nuclease degradation, which can affect the sensitivity of the test if the clinical specimens are not stored properly. Nucleic acid stabilization buffers or Flinders Technology Associates (FTA) cards to transport and store the samples can be used to attenuate these limitations. It should be noted that RNA is a better indicator of active infections than DNA (*35*), making SHERLOCK4HAT a valuable tool for assessing treatment outcome.

The bio-banked clinical samples used here to validate SHERLOCK4HAT did not allow proper analysis of sensitivity for gHAT patients. 44% of confirmed gHAT patient samples were negative with SHERLOCK, and 53% were negative using standard qPCR analysis. The discrepancy between our results compared to the original in-field diagnostic is most probably due to the deterioration of the NA in these samples, that were stored at −80°C for more than 10 years without preservative (*41*). Low parasitemia is typical in gHAT infections, thus any NA degradation could have a dramatic effect on detection using molecular techniques. *T. b. rhodesiense* infections have higher parasitemia, hence deterioration of NA in the samples might have a lower impact in the diagnostic sensitivity, which is evident given the robust sensitivity using rHAT SHERLOCK. The lower sensitivity observed with the *TgSGP* target (42.5%) could be attributed to a selective degradation of the target RNA and / or to a differential expression of the *TgSGP* gene in these samples, since the analytical sensitivity of SHERLOCK for *7SLRNA* and *TgSGP* was similar. From the 62 negative control samples, 7 tested positive for *TgSGP* compared to 1 or 0 for *7SLRNA* SHERLOCK or qPCR, respectively. This reduced specificity needs to be interrogated further.

As we move towards the elimination phase of gHAT, if not to the post-elimination phase in several countries, SHERLOCK4HAT is a viable alternative to currently used diagnostics at the PoC. Optimization of the one-pot reaction to meet the sensitivity requirements for HAT diagnosis, lyophilization of the reaction components and field-friendly NA extraction methods will be required before large-scale deployment. Sensitivity could also be improved using a combination of *7SLRNA, TgSGP* and *SRA* targets in a multiplex SHERLOCK4HAT diagnostic kit that would allow the specific detection of the three subspecies of *T. brucei* at the same time in a single reaction, thereby reducing the full diagnostic cost and making the technology more adapted for horizontal epidemiological studies, including in animal reservoirs. In total, SHERLOCK4HAT provides a readily adaptable diagnostic method for HAT allowing for PoC diagnosis, mass screening and epidemiological surveillance.

## Materials and Methods

### LwCas13a protein expression and purification

Plasmid pC013-Twinstrep-SUMO-huLwCas13a (Addgene plasmid # 90097) (*11*) was used to express LwCas13a in *Escherichia coli* Rosseta™ 2(DE3) pLySs competent cells. Cell pellet was lysed with supplemented lysis buffer (20 mM Tris-HCl pH8.0, 500 mM NaCl, 1 mM DTT, 100 mg lysozyme, 200U Deoxyribonuclease I) and LwCas13a protein was purified from the cleared supernatant as described in (*13*) (fig. S10).

### Target selection and crRNA and RPA primer design

Target genes were selected as either unique or based on their conservation between *Trypanosoma* spp using literature and publicly available data from TriTrypDB (https://tritrypdb.org/tritrypdb/app). Candidate genes were aligned using BLAST with the Trypanosomatidae (taxid:5654) nucleotide collection database from the National Center for Biotechnology Information (NBCI). Alignments to ensure conservation of targets across the *Trypanozoon* subgenus or exclusivity between *T. brucei* subspecies was performed using Clustal Omega. Data available in TriTrypDB was used to identify SNPs found in different field isolates in the target genes *7SLRNA, SODB1* and *TgSGP* (*50*). To identify the SNPs in the *SRA* gene, the sequence variants AF097331, AJ345057, AJ345057 (*22, 26*) were aligned using Clustal Omega and visualized in Jalview 2.11.1. RPA primers and crRNAs were designed to cover the conserved regions of the selected genes and outside of regions containing single nucleotide polymorphisms (SNPs). BLAST analysis with the nucleotide collection of all available genomes was performed to ensure RPA primers and crRNAs specific alignment. A 5’ T7 RNA polymerase promoter sequence (5’GAAATTAATACGACTCACTATAGGG) overhang was added to the RPA forward primers for *in vitro* transcription (IVT) during SHERLOCK reaction. The amplicons generated during RPA reactions are between 130-160 nt length. We used a 28 nt crRNA spacer for all guides in this study except for cr7SLbs which is 26 nt. The spacer sequence is joined to a 5’ direct repeat (DR) to generate the complete crRNA. To facilitate amplification from DNA templates a T7 RNA polymerase promoter sequence was added upstream of the crRNA (spacer + DR + T7 promoter 5’>3’). RPA primer, crRNA and DNA IVT template sequences are detailed in supplementary table 1. SNPs identified for each target gene are included in supplementary table 2.

#### Target RNA and crRNA synthesis and purification

To produce the crRNA’s, DNA IVT templates and T7-3G oligonucleotide were purchased from ThermoFisher. crRNAs were synthesized as described in (*13*) with the following modifications. DNA IVT template (10 µM) and T7-3G oligonucleotide (10 µM) were annealed in standard Taq buffer (1x) by performing a 5-minute denaturation, followed by slow cooling (ramp rate was adjusted to 0.13°C/s) of the reaction to 4°C in a PCR thermocycler (BioRad). IVT was performed using the HiScribe™ T7 Quick High Yield RNA Synthesis Kit (NEB E2050S), where 10 µL of annealed reaction were mixed with 10 µL of NTP buffer mix, 2 µL of T7 RNA polymerase mix and 17 µL of RNase free water. The reaction was incubated for 6 hours at 37°C followed by 15 minutes of DNase I digestion to remove DNA template. Purification of crRNA was done with Agencourt RNAClean XP beads following the manufacturer’s protocol and crRNA concentration was adjusted to 300 ng/µL. Urea Poly-Acrylamid Gel Electrophoresis were used to confirm the purity and correct size of crRNAs.

120 ng of RNA from *T. b. gambiense* ELIANE strain for *TgSGP* or 40 ng of RNA from *T. b. rhodesiense* EATRO strain for *SRA* RNA production were retro-transcribed using pT19 oligonucleotide and SuperScript IV Retro Transcriptase (Thermo Fisher) following standard protocols. cDNAs were purified with Ampure XP (A63880) following manufacturer instructions and eluted in 30 µL of nuclease free water. *TgSGP* and *SRA* were amplified from 5 µL of cDNA, using TgSGP-FL-F and TgSGP-FL-R primers or SRA-FL-F and SRA-FL-R primers, respectively. For 7SLRNA production, the *7SLRNA* gene was amplified from 120 ng of *T. b. brucei* Lister 427 genomic DNA, using 7SLb-UP-F.6 and 7SLb-FL-R primers. The PCR amplification reaction was as follows: 0.2 mM dNTPs, 0.5 µM of each primer, 5 µL of DNA, 0.75 µL of DMSO, 0.5 µL of Phusion DNA polymerase in HF buffer 1X in a 25 µL final volume and was run according to standard PCR settings. A T7 RNA polymerase promoter sequence overhang was included in each forward primer for *in vitro* transcription. IVT of the amplified genes was performed using the HiScribe™ T7 Quick High Yield RNA Synthesis Kit following manufacturer’s instructions and the reaction incubated for 3 hours at 37°C followed by 15 minutes of DNaseI digestion. The single stranded RNA was purified using Agencourt RNAClean XP beads following the manufacturer’s protocol. *In vitro* transcribed RNAs were sequenced and Urea Poly-Acrylamid Gel Electrophoresis were used to confirm the purity and correct size of the target RNAs. Primer sequences used in this section are specified in supplementary table 6.

#### RNA isolation from cultured parasites

*T. b. brucei* Lister 427 bloodstream stage cells were cultured in HMI-11 medium at 37.4°C with 5% CO2. RNA was harvested at 1× 10^6^ cell/mL. *T. b. gambiense* ELIANE and *T. b. rhodesiense* EATRO cell pellets were a gift from Annette MacLeod and *L. major* cells were a gift from Gerald Spaeth. Total RNA from *T. b. brucei* Lister 427, *T. b. gambiense* ELIANE strain, *T. b. rhodesiense* EATRO strain, *L. major* and Human embryonic kidney (HEK) 293T cells, was extracted with the RNeasy Mini kit (Qiagen).

### Simulated samples and total nucleic acid (TNA) extraction from blood, buffy coat and dry blood spots (DBS)

Blood from healthy human donors was provided by ICAReB platform (Clinical Investigation & Access to Research Bioresources) in the Center for Translational Science, at the Institut Pasteur (Paris) (*51*). All participants gave written informed consent in the frame of the healthy volunteers Diagmicoll cohort (Clinical trials NCT 03912246) after approval of the CPP Ile-de-France I Ethics Committee (2009, April 30th). Whole blood was extracted in BD Vacutainer™ Glass ACD Solution Tubes and immediately processed. To determine the performance of *7SLRNA* SHERLOCK in whole blood, buffy coat and DBS, 2×10^5^ *T. b. brucei* Lister 427 parasites were spiked into 20 mL of human blood followed by 1:10 dilution to simulate parasitemia of 10 and 1 parasites/µL. Nine drops of 50 µL of each dilution of the simulated infected blood and non-infected blood were dried on Whatman 903™ Cards and stored at RT for 24 hours until processed. Three tubes of 500 µL of whole blood were snap-frozen and stored at −80°C for 72 hours. To obtain the buffy coat, 12 mL of each simulated infected blood dilution and non-infected blood were centrifuged at 1800 g for 10 minutes without brake, at 4°C to prevent RNA degradation. Three tubes of 125 µL of buffy coat for each dilution were snap-frozen and stored at −80°C for 72 hours. TNA extraction from DBS was performed with the Nucleospin Triprep kit (Macherey-Nagel). For that, 6 × 6 mm punches were incubated with 350 µL of RP1 buffer and 3.5 µL of β-mercaptoethanol at 37°C for 30 minutes in agitation (1000 rpm). The instructions from the manufacturer were followed from this step onwards. DNA and RNA were eluted together in 40 µL of nuclease free water. TNAs from whole blood and buffy coat simulated samples was conducted with Maxwell RSC DNA blood kit (PROMEGA AS1400). The samples were pre-processed with 300 µL of lysis buffer and 30 µL of Proteinase K solution, vortexed for 10 seconds and incubated at 56°C for 20 minutes. The volume was transferred to the Maxwell Cartridge and the kit protocol was run in the automated Maxwell RSC system. TNAs were eluted in s60 µL of elution buffer. Extractions for each dilution and type of sample were done in triplicate.

### Determination of analytical sensitivity

To determine the analytical sensitivity of *7SLRNA* SHERLOCK limiting dilutions of *T. b. brucei* Lister 427 parasites were spiked into un-infected human blood. TNAs were extracted from whole blood and buffy coat with Maxwell RSC DNA blood kit as detailed above. Three replicates of each dilution were assessed by SHERLOCK and the estimated LoD was determined as the lowest concentration where 3 out of 3 replicates were positive. The analytical sensitivity was confirmed in buffy coat by using 20 replicates of 0.66X, 1X and 1.5X the estimated LoD concentration. The analytical sensitivity was determined as the concentration where 95% of the samples gave positive results.

### Optimization of TNA extraction methods

To compare different TNA extraction methods from DBS, sheep blood was spiked with *T. b. brucei* Lister 427 parasites at limiting dilutions (1000-1 parasites/µL). Drops of 50 µL were dried into Whatman 903™ Cards and stored at RT for 24 hours. For TNA extraction with RNeasy mini and micro kits (Qiagen) 3 × 6 mm punches were resuspended with 370 µL of RLT buffer with 3.7 µL of β-mercaptoethanol and incubated at 37°C for 30 minutes with agitation (1000 rpm). The punches and liquid were transferred into a QIAshredder column and spun at maxim speed for 1 minute. The homogenized sample was then processed according to manufacturer’s instructions without DNaseI digestion. DNA and RNA were eluted in the same fraction with 10 µL (RNeasy micro kit) or 30 µL (RNeasy mini kit) of nuclease-free water. For TNA extraction with the Nucleospin Triprep kit (Macherey-Nagel) 3 × 6 mm punches were resuspended with 350 µL of RP1 buffer with 3.5 µL of b-mercaptoethanol and incubated at 37°C for 30 minutes in agitation (1000 rpm). The instructions from the manufacturer were followed from this step onwards and DNA and RNA were eluted together in 40 µL of nuclease free water. Maxwell RSC DNA blood kit (AS1400), Maxwell RSC SimplyRNA blood kit (AS1380) and RNeasy mini kit (Qiagen) were used to extract TNA from 250 µL or 125 µL of human buffy coat spiked with limiting dilutions of *T. b. brucei* Lister 427 parasites.

### Field isolated samples

The RNA used in this study was derived from 57 field isolates representing different *Trypanosoma* species, subspecies and strains. The collection contained 50 *T. b. gambiense* group 1 (46 bloodstream forms + 4 insect forms), 1 *T. b. gambiense* group 2, 2 *T. b. rhodesiense*, 1 *T. b. brucei*, 1 *T. equiperdum* and 2 *T. evansi* strains or clones. They were kindly provided by Nick Van Reet and Philippe Büscher (Institute of Tropical Medicine [ITM], Antwerp, Belgium) (*40*). The RNA was kept at −80 and the concentration normalized to 5 ng/µL. Three microliters of input material were used for each SHERLOCK analysis.

### Clinical samples

Clinical samples in this study were obtained from the WHO HAT Specimen biobank (*41*). They included buffy coats from 48 individuals living in *T. b. gambiense* endemic areas who were negative for gHAT by serology and parasitology, 50 patients with confirmed gHAT at stage 1, 48 patients with confirmed gHAT at stage 2, 20 individuals living in *T. b. rhodesiense* endemic areas who were negative for rHAT by serology and parasitology and 19 patients with confirmed rHAT at stage 2. TNA were extracted from 125 µL of sample using the Maxwell RSC Blood DNA kit as detailed above. The material was eluted in 50 µL of elution buffer.

### SHERLOCK two-step reaction

For the isothermal amplification step, TwistAmp Basic kit (TwistDx) was used according to manufacturer’s instructions with the following modifications. For each reaction, 3 µL of input total nucleic acids (TNA) were incubated with 480 nM of each RPA primer (240 nM for *TgSGP* RPA primers), Reaction buffer 1X, 2.2 U of Transcriptor (Roche), 1.5 U of Murine RNase inhibitor (NEB) and 14 mM MgOAc and 0.22 pellet of TwistAmp Basic kit, in a final volume of 11 µL. Reaction condition were optimized using different RPA primer concentrations (120, 240 or 480 nM) and MgOAc concentrations (14, 22 or 30 mM). The reactions were run using Hard-shell thin wall 96 well PCR Plates, sealed with Microseal ‘F’ Foil Seals (BioRad). Plates were incubated in a heating block set to 42°C with thermoregulated lid. After a 5 minutes incubation, the plates were agitated for 15 seconds and the incubation resumed for 40 min. For the LwCas13a detection step, 1 µL of the previous reaction was incubated with 20 mM HEPES pH 6.5, 9 mM MgCl2, 1 mM rNTP mix (NEB), 126 ng of LwCas13a, 2 U of Murine RNase inhibitor (NEB), 25 U of NxGen T7 RNA Polymerase (Biosearch technology), 10 ng of crRNA and 125 nM of RNaseAlert probe V2 (Invitrogen) in a final volume of 20 µL. The reactions were run in 3 or 4 replicates in 384-well plates, F-bottom, µClear bottom, black, sterile, with lid (Greiner). The incubations were maintained at 37°C in the TECAN plate reader INFINITE 200 PRO Option M PLEX and the fluorescence was recorded at an initial time point and after 2 h 30 min or 3 hours. For the lateral flow assay (LFA) readout Milenia HybriDetect strips were used and the RNaseAlert probe was substituted for 10 pmol of LF-RNA reporter (/56-FAM/rUrUrUrUrUrU/3Bio) and was incubated likewise. Following this, the SHERLOCK reaction was mixed with 80 µL of a PEG-based CRISPR-optimized Lateral Flow Assay Buffer (provided by Milenia Biotec GmbH, Germany). The strip was dipped in the mix and the results were interpreted after 5 min.

### SHERLOCK one-step reaction

For the single step SHERLOCK assay, 8 µL of input NA were mixed with 1 pellet of TwistAmp Basic kit, 20 mM HEPES pH 8, 60 mM KCl, 5% PEG-8000, 132 ng of LwCas13a in 1mM Tris-HCl pH 7.5, 12 mM NaCl, 0.1% glycerol, 125 nM of RNaseAlert probe, 2 U/µL of ProtoScript II RT (NEB), 0.1 U/µL of RNase H (NEB), 1 U/µL of NxGen T7 RNA Polymerase (Biosearch technology), 455 nM of each RPA primer, 10 nM of crRNA and 14 mM of MgOAc in a final volume of 107.5 µL. For each technical replicate, 20 µL of the mix were transferred to a 384-well plate, F-bottom, µClear bottom, black, sterile, with lid (Greiner). The incubations were done in the TECAN plate reader as described above. The fluorescence was monitored over 2 h 30 min at 37°C with a 30 min interval between acquisitions.

### Quantitative PCR analysis

TNA were analyzed by qPCR using Luna Universal qPCR MasterMix (NEB). The qPCR amplification mix contained 1 µL template and 0.4 µM of each primer (Tb177bp F/R). Reactions were run in triplicate in a Hard-shell PCR Plates 96 well, thin wall, which were sealed with Microseal ‘B’ Seals (BioRad). All experiments were run on a CFX96 Touch Real-time Detection system with a C1000 Touch Thermal cycler (Bio-Rad), using the following PCR cycling conditions: 50°C for 5 min, 95°C for 10 min, then 40 cycles of 95°C for 15 sec and 66°C for 1LJmin (fluorescence intensity data collected at the end of the last step), followed by a temperature gradient between 66°C and 95°C. The last step was used for dissociation analysis of the PCR product to monitor the amplicon identity. For that, the melt temperature of the amplicons from clinical samples was compared with that from *T. brucei* control nucleic acids. Sequence of primers in this section are listed in supplementary table 6.

### Data analysis

We used the fluorescence given by the negative control, where water is used as input material, as the background fluorescence. To calculate the background-subtracted fluorescence intensities in a given multi-well plate, we subtracted the background fluorescence from each sample fluorescence at final time point. To calculate the fold-change over background fluorescence in a given multi-well plate, sample fluorescence was divided by background fluorescence at final time point. For the optimization of the TNA extraction methods, fluorescence values were reported as fold-changes from the initial baseline fluorescence intensity by dividing the fluorescence value at last time point by the value at initial time point. Baseline initial fluorescence and background fluorescence differ between runs. Therefore, it was more effective to compare fold-change over background fluorescence.

For the analysis of the clinical samples the following ratios were calculated for every target assessed

- Negative template controls Ratio (Rntc) = Fold-change over the initial baseline fluorescence

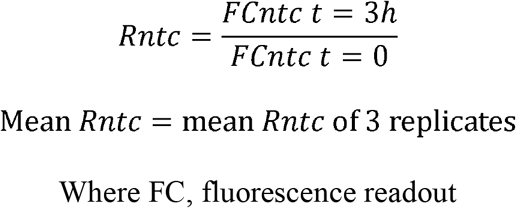
- Positive template controls Ratio (Rpc) and sample Ratio (Rsample) = Fold-change over the background fluorescence at time 3h. Background fluorescence is given by the negative template control reaction

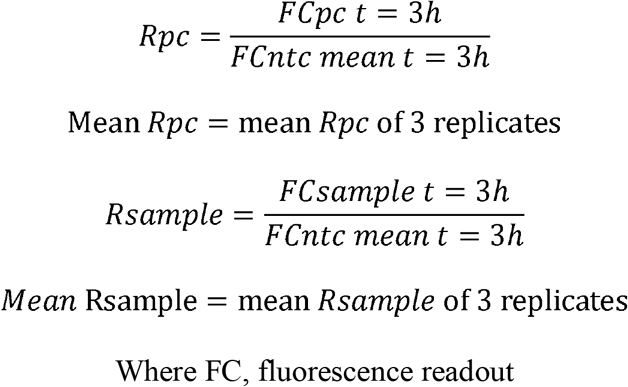

Fluorescence ratios from positive and negative samples were computed by receiver operating characteristic (ROC) curve analysis for determining test positivity thresholds (fig. S11).

For qPCR, mean Ct values of 3 technical replicates were reported. A read was considered positive when the Ct value was lower than the established cut-off and the identity of the amplicon of these assays was confirmed by dissociation analysis (specific melt temperature ± 0.5 degrees). The specific melt temperature for each amplicon was calculated by dissociation analysis of the qPCR amplicons using dilution series of TNA from cultured parasites. The Ct value cut-off was determined using cumulative distribution analysis (fig. S12).

All plots and statistical analyses were performed with GraphPad Prism 9.1.2.

## Data Availability

All data are available in the main text or the supplementary materials.

## Acknowledgments

We would like to thank the following people for kindly providing parasite strains and / or parasite NAs used in this study: Annette MacLeod (University of Glasgow, Glasgow, UK), Gerald Spaeth and Artur Scherf (Institut Pasteur, Paris, France), Philippe Büscher and Nick Van Reet (Institute of Tropical Medicine, Antwerp, Belgium).

## Author contributions

NS, BR and LG conceived and designed the experiments. NS, ADH performed the experiments. NS, BR and LG analyzed the data. MNU, BLP, BR and LG contributed reagents, materials and analysis tools. NS, BR and LG wrote the paper.

## Data and materials availability

All data are available in the main text or the supplementary materials.

## Ethics statement

Blood from healthy human donors was provided by ICAReB platform (Clinical Investigation & Access to Research Bioresources) in the Center for Translational Science, at the Institut Pasteur (Paris). All participants gave written informed consent in the frame of the healthy volunteers Diagmicoll cohort (Clinical trials NCT 03912246) after approval of the CPP Ile-de-France I Ethics Committee (2009, April 30th). For the WHO HAT Specimen biobank samples, approval was given by the WHO Ethical Review Committee, each national ethical committee where samples were taken and the national Ministries of Health.

