## Supplementary material for "SHERLOCK4HAT: a CRISPR-based tool kit for diagnosis of Human African Trypanosomiasis": Fig. S

**This PDF file includes:**

Figs. S1 to S12
Tables S2 to S3 and S6
Legends for tables S1, S4, S5 and data files S1 to S2

**Other Supplementary Materials for this manuscript include the following:**

Tables S1, S4 and S5
Data files S1 to S2

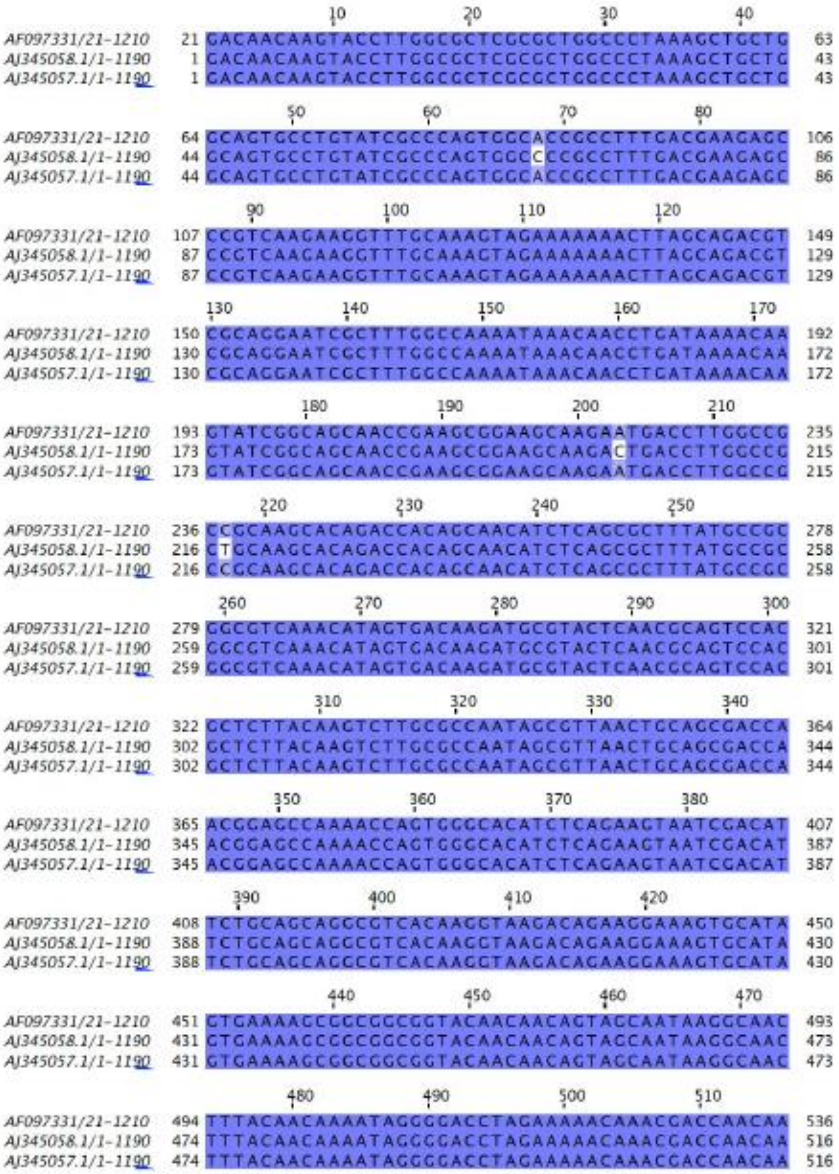
AF097331/21-1210
AJ345058.1/1-1190
AJ345057.1/1-1190

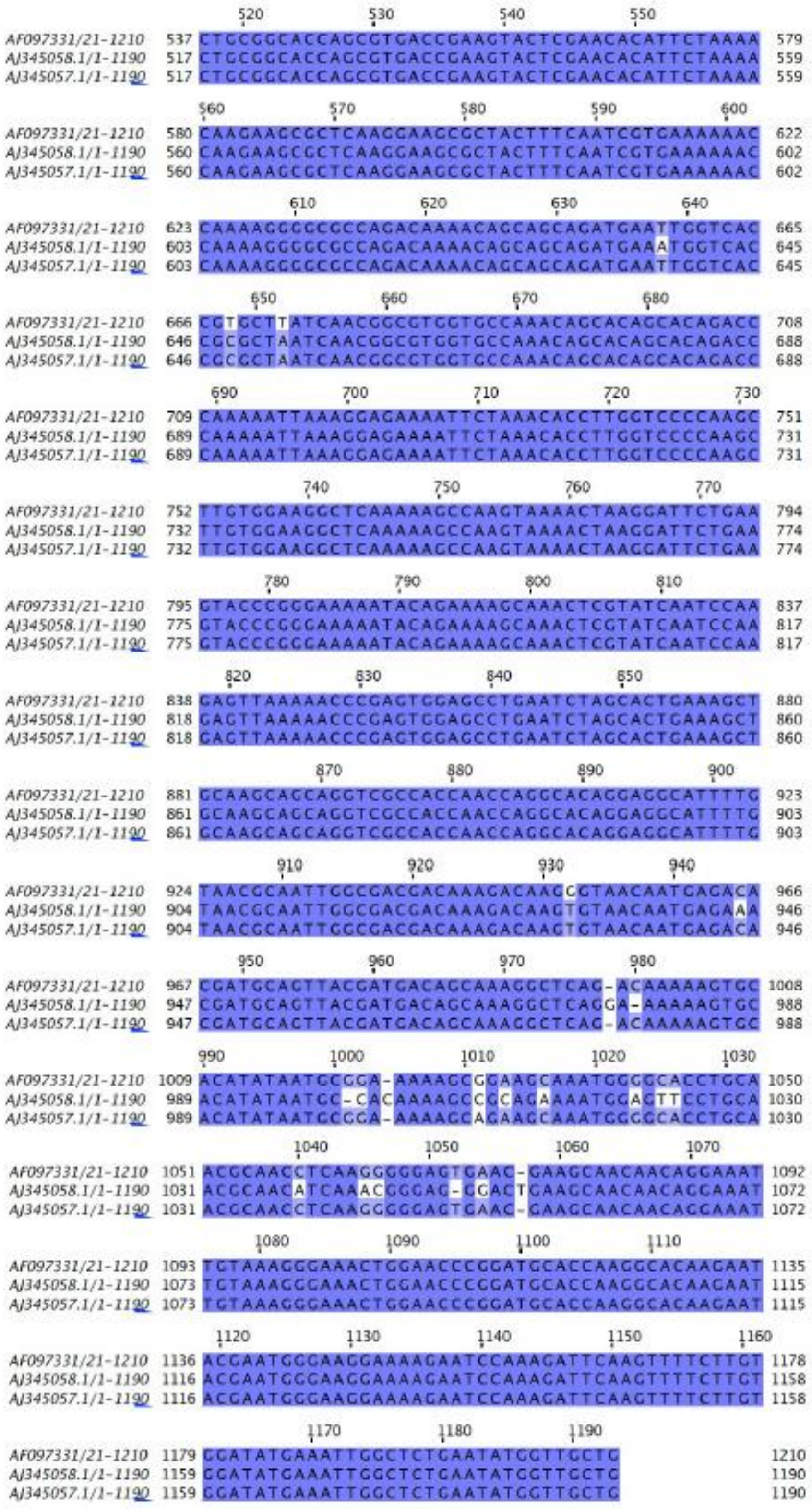

**Fig. S1. Comparison of *SRA* sequences AF097331, AJ345058.1 and AJ345057.1.**

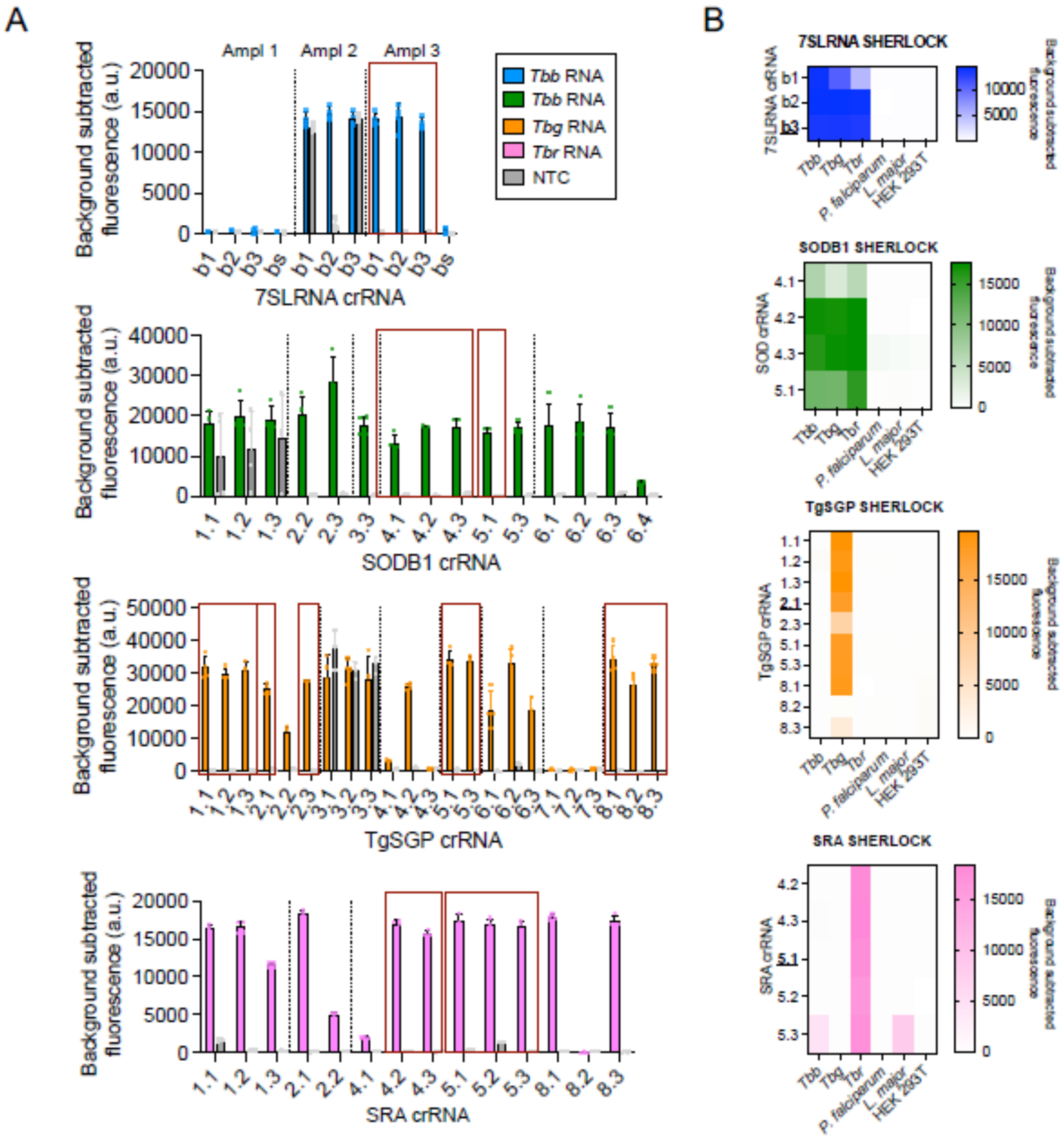


**Fig. S2. RPA primer pair and CRISPR RNA guide screening for *Trypanosoma bruce*i spp. target genes. (A),** SHERLOCK performance using multiple combinations of RPA primer pairs and crRNA covering *7SLRNA*, *SODB1*, *TgSGP* and *SRA* gene sequences. Bars represent the mean background subtracted fluorescence of 4 technical replicates + SD. Total RNA at 5 ng/µL from *T. b. b.* Lister 427, *T. b. g.* ELIANE strain or *T. b. r.* EATRO strain was used for the *7SLRNA* and *SODB1*, *TgSGP* and *SRA* reactions, respectively. Red boxes highlight the crRNA that were further analysed. **(B),** Specificity of selected RPA primers and crRNA guides was assessed using RNA from *T. b. b.* Lister 427, *T. b. g.* ELIANE strain, *T. b. r.* EATRO strain, *P. falciparum*, *L. major* and human embryonic kidney (HEK) 293T cells. In bolt, the selected guides for following experiments. NTC, non-template control; *Tbb*, *T. b. brucei* ; *Tbg*, *T. b. gambiense* ; *Tbr*, *T. b. rhodesiense*.

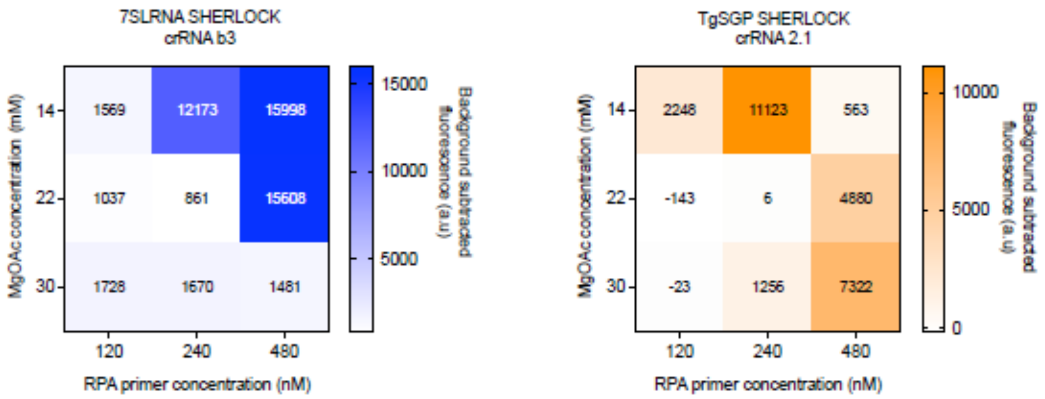


**Fig. S3. Heat maps of RPA primer and MgOAc concentration combinations to detect *7SLRNA* and *TgSGP* targets.** RPA screens comprised of 9 set combinations of 3 MgOAc and 3 primer pair concentrations for best-performing condition to amplify sequence targets. Input ssRNA at 2000 aM for *7SLRNA* and 200 aM for *TgSGP*.

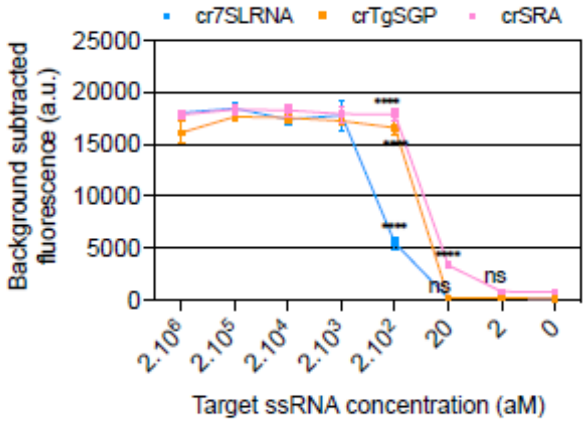


**Fig. S4. SHERLOCK limit of detection (LoD) in the attomolar range.** Dilution series of *in vitro* transcribed target *7SLRNA*, *TgSGP* and *SRA* RNAs were used for the *7SLRNA*, *TgSGP* and *SRA* SHERLOCK reactions respectively. Student's two-tailed t test between fluorescence output of sample versus no-template control. **** $p<0.0001$.

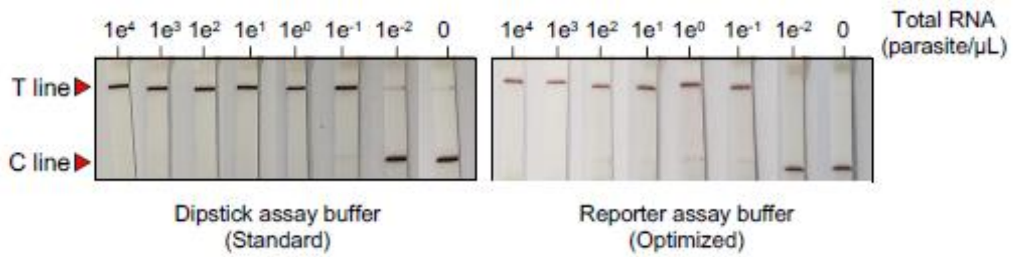


**Fig. S5. Lateral Flow Assay (LFA) optimization.** Performance of two LFA buffers. A reporter assay buffer or a dipstick assay buffer were used to dilute the *7SLRNA* SHERLOCK reaction prior the lateral flow assay. The signal was read after 5 minutes of incubation. Left panel is the experiment shown in figure 1E for comparison.

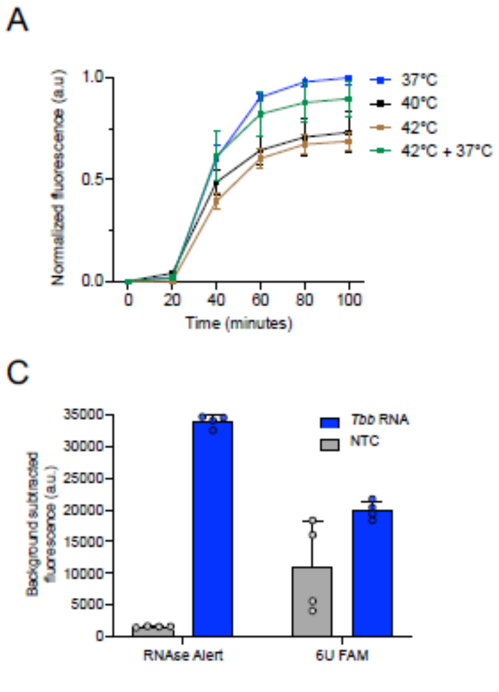


**Fig. S6. Optimization of one-tube reaction. (A),** Comparison of different incubation temperatures. Three different temperatures were assessed, 37°C, 40°C and 42°C, and a combination of 42°C for 10 minutes (optimal for RT-PCR) followed by 37°C for the rest of the reaction (optimal for Cas13a). Incubation at 37°C resulted to be the best performance temperature, in terms of speed and signal. RNA from 100 parasite/µL was used in each reaction. **(B),** Retro transcriptase enzymes from different manufacturers were compared. ProtoScript II from NEB was selected for the optimized reaction as it gave higher signal and was more cost-effective. RNA from 100 parasite/µL was used in each reaction. **(C),** Two RNA quenched fluorescent RNA reporters were tested. RNase Alert was selected for the optimized reaction as it led to a higher signal-to-noise ratio. RNA from 100 parasite/µL was used in each reaction. **(D),** Different input volumes were assessed in the one-tube reaction. For the optimized reaction 8 µL of input volume was used to increase the sensitivity of the test.

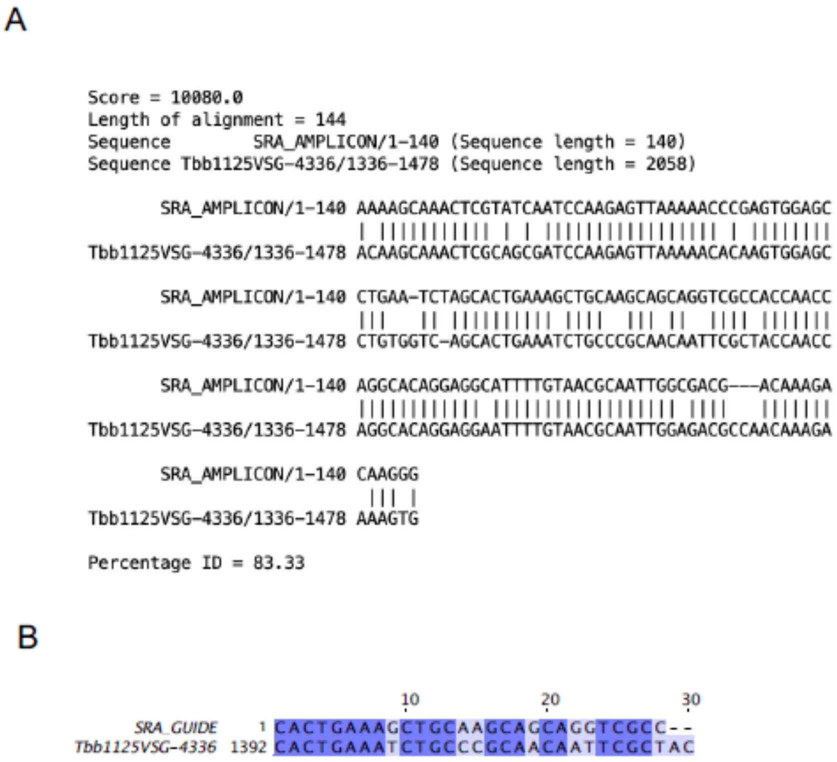


**Fig. S7. Alignment of *SRA* amplicon and guide with Tbb1125VSG-4336. (A),** Clustal alignment of *SRA* SHERLOCK amplicon with Tbb1125VSG-4336 expressed in the triple positive field isolated sample AnTat 22.1. **(B),** Nucleotide mismatches between the *SRA* crRNA and Tbb1125VSF-4336.

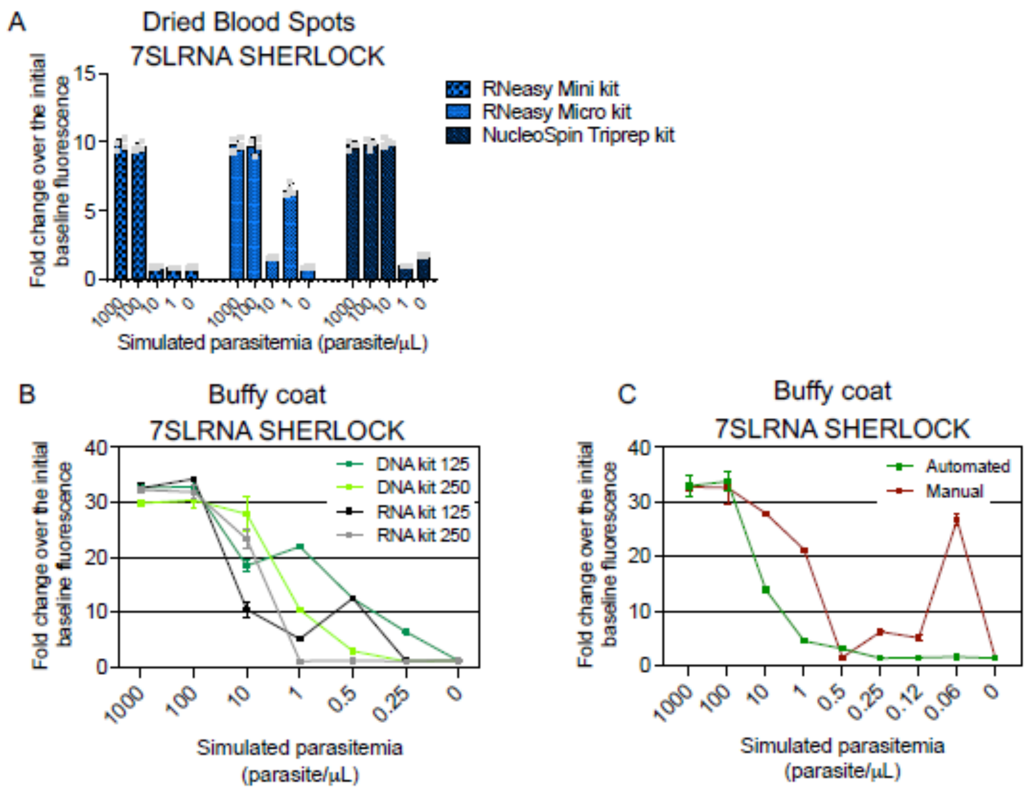


**Fig. S8. Total nucleic acid extraction optimization. (A),** Comparison of TNA extraction from DBS. Fifty microliters of spiked sheep blood was dried on Whatman 903™ Cards and stored for 24 hours at room temperature. TNA were extracted from 3 punches of 6mm using the RNeasy mini kit (QIAGEN), RNeasy micro kit (QIAGEN) or the NucleoSpin Triprep (Macherey-Nagel). Bars represent mean readout +SD of 4 SHERLOCK replicates, shown as grey circles. **(B),** Trypanosome TNAs extraction from spiked human buffy coat using Maxwell automated system. Two Maxwell RSC kits (simplyRNA blood kit and Blood DNA kit) and two different input volumes (125 µL and 250 µL) were compared. **(C),** SHERLOCK assay for *7SLRNA* target. Uninfected human blood was spiked with 1000 p/µL followed by dilution series. Buffy coat was obtained by centrifugation and TNAs were purified either with Maxwell automated system (Maxwell RSC Blood DNA kit) or with the manual column-based QIAGEN RNeasy mini kit, with minor modifications (see materials and methods). The data from the Maxwell extraction system is also shown in figure 3 B.

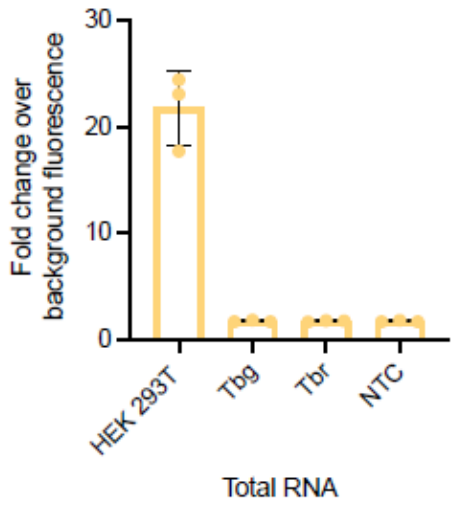


**Fig. S9. Human *RNase* P SHERLOCK assay.** Total RNA from human cells (1ng/µL) or cultured parasites (5ng/µL) was assessed with *RNase P* SHERLOCK. *RNase P* test performed well and showed no cross-reactivity with parasite RNA. Bars represent the mean of three technical replicates ±SD.

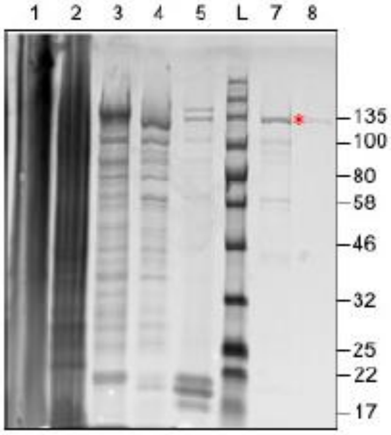


**Fig. S10. LwaCas13a protein expression and purification.** Protein fractions collected during purification were analyzed by SDS-PAGE followed by Coomassie Blue staining. 1, cell pellet after clearing cell lysate; 2, cleared cell lysate supernatant; 3, flow-through following Strep-Tactin resin binding; 4, Strep-Tactin resin after SUMO protease cleavage; 5, eluted fraction post SUMO protease cleavage; L: Ladder; 7, eluted fraction after ion exchange chromatography (HiTrap column); 8, final product after size exclusion chromatography (Superdex column). The red * at 135 kDa indicates the band corresponding to the purified LwaCas13a protein used in this study.

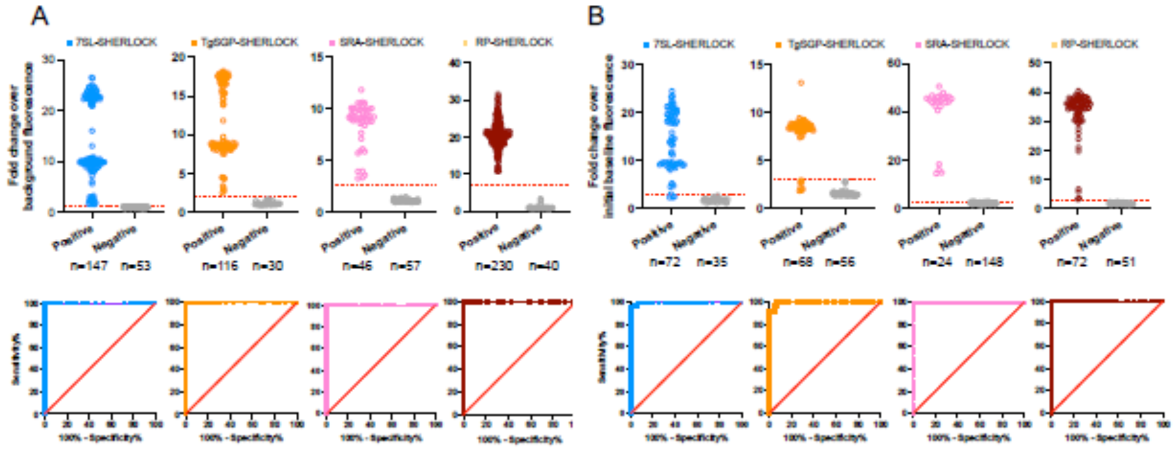


C

| | Threshold for positivity | | | | |
|---|---|---|---|---|---|
| | Threshold | Sensitivity (%) | 99% CI | Specificity% | 99% CI |
| 7SL-SHERLOCK | > 1.290 | 100 | 95,68% to 100,0% | 100 | 88,87% to 100,0% |
| TgSGP-SHERLOCK | > 2.065 | 100 | 94,59% to 100,0% | 100 | 81,89% to 100,0% |
| SRA-SHERLOCK | > 2.320 | 100 | 87,39% to 100,0% | 100 | 89,57% to 100,0% |
| RP-SHERLOCK | > 7.145 | 100 | 97,20% to 100,0% | 100 | 85,77% to 100,0% |

| | Threshold for assay validation | | | | |
|---|---|---|---|---|---|
| | Threshold | Sensitivity (%) | 99% CI | Specificity% | 99% CI |
| 7SL-SHERLOCK | > 2.925 | 95,83 | 84,99% to 99,39% | 100 | 84,06% to 100,0% |
| TgSGP-SHERLOCK | > 2.875 | 91,18 | 78,30% to 96,73% | 100 | 89,41% to 100,0% |
| SRA-SHERLOCK | > 2.936 | 100 | 78,34% to 100,0% | 99,32 | 94,50% to 99,99% |
| RP-SHERLOCK | > 2.885 | 100 | 91,56% to 100,0% | 100 | 88,49% to 100,0% |

**Fig. S11. Receiver operating characteristic (ROC) curve analyses of positive and negative samples and threshold ratio calculation. (A),** Laboratory generated positive and negative sample SHERLOCK ratios (fold change over background fluorescence) and ROC curves for *7SLRNA*, *TgSGP*, *SRA* and *RNase P* targets to calculate threshold for positivity. **(B),** Laboratory generated positive and negative sample SHERLOCK ratios (fold change over initial fluorescence) and ROC curves for *7SLRNA*, *TgSGP*, *SRA* and *RNase P* targets to calculate threshold ratio for assay validation. **(C),** Tables summarizing threshold ratios for each target and estimated sensitivity and specificity with the 99% CI.

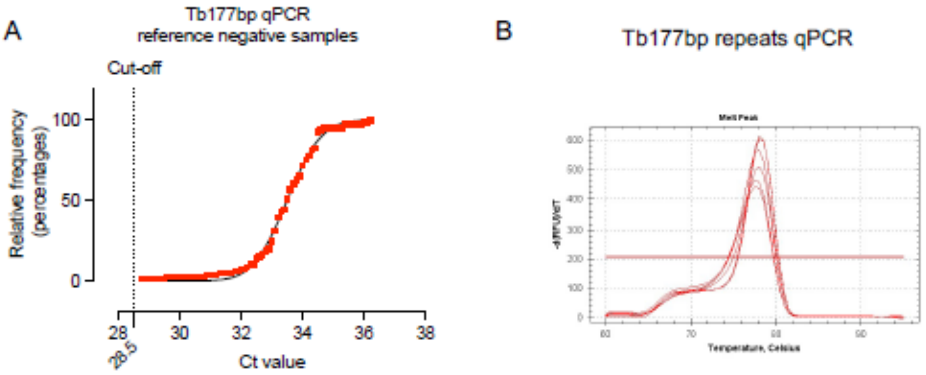


**Fig. S12. Criteria for validation qPCR reads. (A),** Cumulative distribution of the Ct values from Tb177bp qPCR of 82 reference negative samples to stablish the Ct value cut-off. Samples were considered positive when the identity of the amplicon is been confirmed by dissociation analysis (specific melt temperature ±0.5 degrees). **(B),** Dissociation analysis of the qPCR amplicons from a dilution series of TNA using cultured parasites. The specific melt temperature for the Tb177bp repeats amplicon is 78.

**Table S2. Single nucleotide polymorphisms identified in the genes of this work.**

| Gene name | SNP ID | Position | REF strain 927 | Major Allele | Minor Allele | Ref. |
|---|---|---|---|---|---|---|
| *7SLRNA* | NGS_SNP.Tb927_08_v5.1.861102 | 155 | T | T (.99) | C (.01) | (50) |
| | NGS_SNP.Tb927_08_v5.1.861053 | 204 | G | A (.91) | G (.09) | |
| | NGS_SNP.Tb927_08_v5.1.861025 | 232 | A | G (.97) | A (.03) | |
| *TgSGP* | | 277 | A | | C | (16) |
| | | 360 | GC | | CG | |
| | | 735 | AT | | TA | |
| | | 797 | GC | | CG | |
| | | 874 | G | | A | |
| *SRA* | | 88, 223, 237, 658, 668, 672, 952, 965, 998, 999, 1020, 1021, 1022, 1029, 1031, 1034, 1041, 1043, 1044, 1058, 1063, 1064, 1070, 1073, 1074 | | | | (22) |
| *SODB1* | NGS_SNP.Tb927_11_v5.1.4212379 | 128 | T | T (.99) | C (.01) | (50) |
| | NGS_SNP.Tb927_11_v5.1.4212294 | 213 | G | A (.86) | G (.14) | |
| | NGS_SNP.Tb927_11_v5.1.4212291 | 216 | A | G (.85) | A (.15) | |
| | NGS_SNP.Tb927_11_v5.1.4212279 | 228 | C | T (.85) | C (.15) | |
| | NGS_SNP.Tb927_11_v5.1.4212278 | 229 | G | A (.85) | G (.15) | |
| | NGS_SNP.Tb927_11_v5.1.4212089 | 418 | T | T (.56) | G (.44) | |
| | NGS_SNP.Tb927_11_v5.1.4211938 | 569 | A | A (.99) | G (.01) | |

**Table S3. SHERLOCK performance of the screened crRNAs.** Background subtracted fluorescence (a. u.) ≤ 1500, no signal; Background subtracted fluorescence (a. u.) >1500 and ≤ 5000, Low; Background subtracted fluorescence (a. u.) > 5000 and ≤ 15000, Medium; Background subtracted fluorescence (a. u.) >15000, High; NA, not assessed; Tbb, *T. b. bucei*; Tbg, *T. b. gambiense*; Tbr, *T. b. rhodesiense*; sspp, subspecies.

| Target | crRNA name | SHERLOCK performance | SHERLOCK specificity |
|---|---|---|---|
| 7SLRNA | cr7SLbs | No signal | NA |
| | cr7SLb1 | High (Tbb) Medium (Tbg) Low (Tbr) | Tb sspp |
| | cr7SLb2 | High | Tb sspp |
| | cr7SLb3 | High | Tb sspp |
| TgSGP | crSGP1.1 | High | Tbg |
| | crSGP1.2 | High | Tbg |
| | crSGP1.3 | High | Tbg |
| | crSGP2.1 | High | Tbg |
| | crSGP2.2 | Medium | NA |
| | crSGP2.3 | High | Tbg |
| | crSGP3.1 | Background | NA |
| | crSGP3.2 | Background | NA |
| | crSGP3.3 | Background | NA |
| | crSGP4.1 | Low | NA |
| | crSGP4.2 | High | NA |
| | crSGP4.3 | No signal | NA |
| | crSGP5.1 | High | Tbg |
| | crSGP5.3 | High | Tbg |
| | crSGP6.1 | High | NA |
| | crSGP6.2 | High | NA |
| | crSGP6.3 | High | NA |
| | crSGP7.1 | No signal | NA |
| | crSGP7.2 | No signal | NA |
| | crSGP7.3 | No signal | NA |
| | crSGP8.1 | High | Tbg |
| | crSGP8.2 | High | Tbg |
| | crSGP8.3 | High | Tbg |
| SRA | crSRA1.1 | High | NA |
| | crSRA1.2 | High | NA |
| | crSRA1.3 | Medium | NA |
| | crSRA2.1 | High | NA |
| | crSRA2.2 | Low | NA |
| | crSRA4.1 | Low | NA |
| | crSRA4.2 | High | Tbr |
| | crSRA4.3 | High | Tbr |
| | crSRA5.1 | High | Tbr |
| | crSRA5.2 | High | Tbr |
| | crSRA5.3 | High | Tbb, Tbr, L. major |
| | crSRA8.1 | High | NA |
| | crSRA8.2 | No signal | NA |
| | crSRA8.3 | High | NA |
| SODB1 | crSOD1.1 | Background | NA |
| | crSOD1.2 | Background | NA |
| | crSOD1.3 | Background | NA |
| | crSOD2.2 | High | NA |
| | crSOD2.3 | High | NA |
| | crSOD3.3 | High | NA |
| | crSOD4.1 | High | Tb sspp |
| | crSOD4.2 | High | Tb sspp |
| | crSOD4.3 | High | Tb sspp |
| | crSOD5.1 | High | Tb sspp |
| | crSOD5.3 | High | NA |
| | crSOD6.1 | High | NA |
| | crSOD6.2 | High | NA |
| | crSOD6.3 | High | NA |
| | crSOD6.4 | Low | NA |

**Table S6. Other oligonucleotides.**

| Name | Sequence 5'>3' | Ref. |
|---|---|---|
| T7-3G | GAAATTAATACGACTCACTATAGGG | *(13)* |
| 7SLb UP F.6 | GAAATTAATACGACTCACTATAGGGAGCCGGAGCGCATT GCTCTGTAACC | This study |
| 7SLb FL R | CCGCCTCGCGACGACACTTGGGGCGCAAA | This study |
| TgSGP FL F | GAAATTAATACGACTCACTATAGGGATGTGGCAATTACT AGCAATAGCGGCGG | This study |
| TgSGP FL R | TTAAAAAAGCAAAAATGCAAGCAAAAGAGGGGCC | This study |
| SRA FL F | GAAATTAATACGACTCACTATAGGGATGCCCCGAAATTC GGGCCGGACAACAA | This study |
| SRA FL R | TTAAAACAGAAAGGCCACAAAAGCAGCAA | This study |
| Tb177F | AACAATGCGCAGTTAACGCTAT | *(32)* |
| Tb177R | ACATTAAACACTAAAGAACAGCGTTG | *(32)* |
| 6U FAM | 56-FAM/rUrUrUrUrUrU/3IABkFQ | *(38)* |

**Table S1. (separate file)**
**RPA primers and crRNA templates.** Selected RPA primer pairs and crRNAs for each target gene are highlighted.

**Table S4. (separate file)**
**Field isolated samples classification and SHERLOCK analysis.** BSF, bloodstream form; PCF, procyclic form; DRC, Democratic Republic of Congo; ITM, Institute of Tropical Medicine, Antwerp, Belgium; NA, (information) not available.

**Table S5. (separate file)**
**Concordance tables, sensitivity and specificity of SHERLOCK4HAT diagnostic kit for detection of trypanosome nucleic acids in buffy coat samples.**

**Data file S1. (separate file)**
**BLAST hits of *7SLRNA* and *SODB1* sequences.** Hits obtained running the *7SLRNA* (Tb927.82861) and *SODB1* (Tb927.11.15910) sequences on the *Trypanozoon* nucleotide data base using BLASTn from the National Center for Biotechnology Information (NBCI).

**Data file S2. (separate file)**
**WHO HAT specimen biobank clinical samples.** Individual information, SHERLOCK4HAT and qPCR results.
